# Age-Specific All-Cause Mortality Trends in the UK: Pre-Pandemic Increases and the Complex Impact of COVID-19

**DOI:** 10.1101/2024.07.14.24310375

**Authors:** Francisco J. Pérez-Reche

## Abstract

**Objectives:** This study aims to analyse age-specific all-cause mortality trends in the UK before, during, and after the COVID-19 pandemic to determine if pre-pandemic trends contributed to increased mortality levels in the post-pandemic era.

**Methods:** We utilised age-structured population and mortality data for all UK countries from 2005 to 2023. Mortality rates were calculated for each age group, and excess mortality was estimated using the Office for National Statistics (ONS) method.

**Results:** Our most concerning finding is an increase in all-cause mortality rates for middle-aged adults (30-54 years) starting around 2012. The COVID-19 pandemic may have further impacted these rates, but the pre-existing upward trend suggests that current elevated mortality rates might have been reached regardless of the pandemic. This finding is more alarming than the slowdown in the decline of cardiovascular disease death rates for individuals under 75 noted by the British Heart Foundation.

**Conclusion:** Our results highlight the importance of considering both immediate pandemic impacts and long-term mortality trends in public health strategies. This underscores the need for targeted interventions and improved healthcare planning to address both ongoing and future challenges.

## Introduction

Excess mortality was observed globally during the COVID-19 pandemic^1–3^. Following the pandemic, many countries, including the UK, continued to experience higher-than-expected mortality levels^4–7^. Reports have highlighted increases in deaths from cardiovascular diseases, cancer, and mental health-related conditions. Contributing factors include healthcare backlogs, delayed diagnoses and treatments of chronic conditions, and a mental health crisis exacerbated by the pandemic. Particularly concerning are the excess deaths among middle-aged adults, which have been attributed to conditions such as cardiovascular disease and cancer ^5,6^.

Since the COVID-19 pandemic, most analyses of UK mortality have concentrated on the pandemic and post-pandemic periods. Mortality trends before the pandemic have received less attention, despite reports of deterioration in some causes of death. Notably, mortality rates for individuals under 75 associated with cardiovascular diseases were steadily improving until 2012, after which the rate of improvement slowed, and post-pandemic rates have remained worryingly high^6^.

In this study, we examine age-specific mortality trends before the COVID-19 pandemic, which are crucial for interpreting the current situation.

## Methods

Our analyses utilise age-structured population and mortality data for all UK countries between 2005 and 2023. The data is used as provided by Ref. ^8^ from the Office for National Statistics, National Records of Scotland, and the Northern Ireland Statistics and Research Agency. Data for England & Wales was pooled into a single group.

Mortality trends were studied using mortality rates and excess mortality. For a given age group, the mortality rate is calculated by dividing the observed deaths in that group by the population of the group. This can be interpreted as the probability that an individual within this age group will die during the considered period. To quantify the uncertainty of the mortality rate, we interpret it as a binomial probability and use “exact” Clopper-Pearson confidence intervals^9^.

The trends in mortality rates between 2012 and 2019 were modelled using linear regression. These models were then used to predict post-2019 trends, allowing us to compare these predictions with the observed trends. This comparison helps assess the impact of COVID-19 and determine whether the mortality trends observed in 2023 could have been reached regardless of the pandemic.

Excess mortality measures the number of deaths in a population during a crisis that exceeds what would be expected under normal conditions. Among the numerous methods to estimate excess deaths^1–3^, we have chosen the new method developed by the Office for National Statistics (ONS)^8^.

## Results

### Mortality rates

Four age groups can be identified in terms of the mortality rate trajectories in the UK between 2005 and 2023: A group of younger individuals that are less than 30 years old, a group of middle-aged adults with ages between 30 and 54 years, a group of older adults aged between 55 and 89 years, and a group of those aged 90 and above. The mortality rate for these groups are illustrated in the plots of the left column of Figure 1. The trends for all the age strata are provided in the Figures 1-15 of Appendix A.

**Figure 1.**
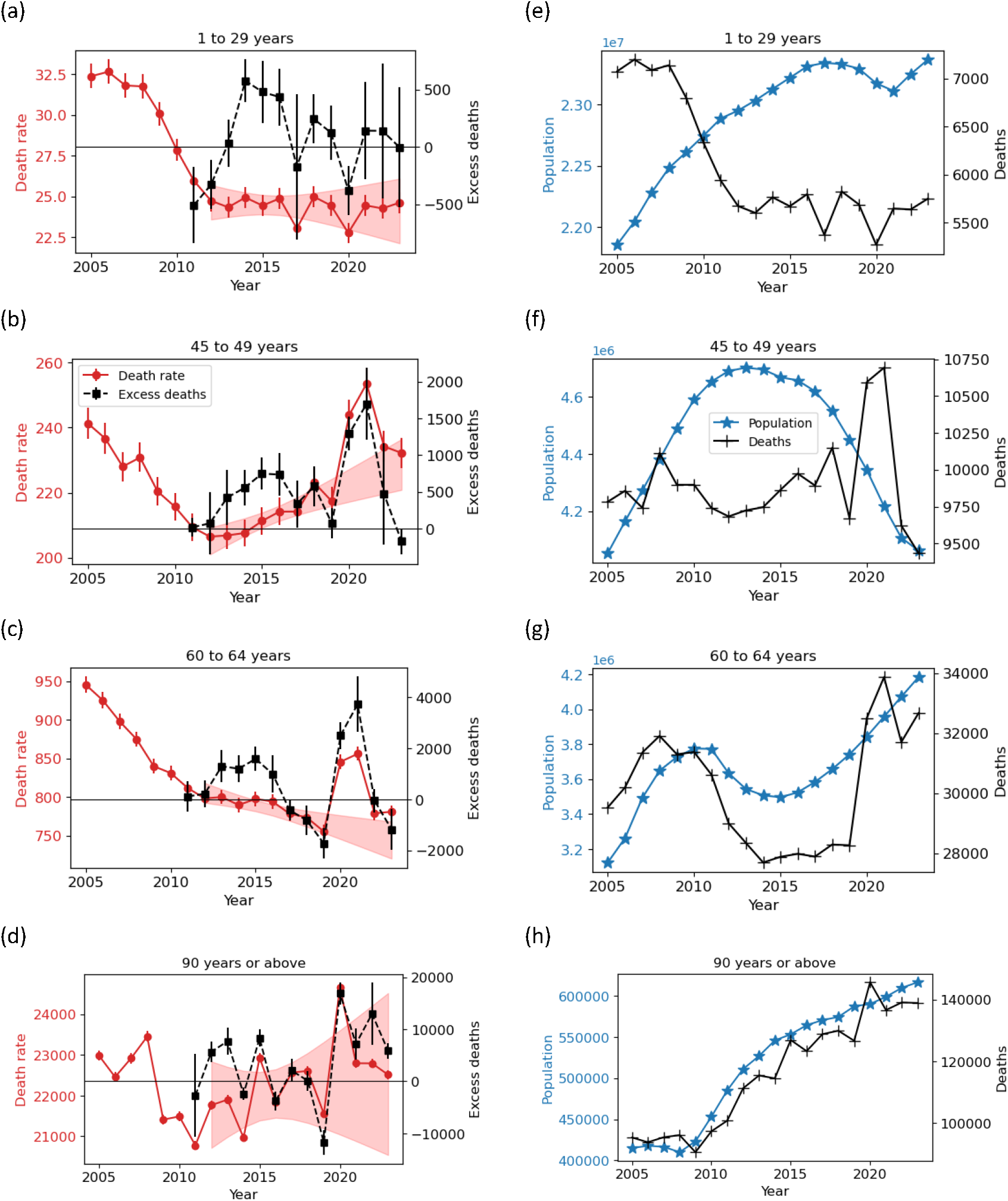
Death rate (per 100,000), excess deaths, population, and registered deaths in the UK for age groups as follows: (a,e) 1 to 29 years, (b,f) 45 to 49 years, (c,g) 60 to 64 years, and (d,h) 90 years or above. Vertical error bars indicate 95% confidence intervals for death rate and excess mortality data. Shaded regions represent 95% confidence intervals for mortality rate predictions derived from linear model fitting on data spanning 2012 to 2019. Analogous results for more age groups are provided in Appendix A.

For ages between birth and 89 years, mortality rates fell until 2012, likely due to improvements in healthcare. Trends after 2012 varied by age group.

For individuals below 30 years old, death rates remained largely constant from 2012 to 2023 (see Figure 1(a)). The lowest rate was recorded in 2020 but no clear trend associated with the COVID-19 pandemic can be identified for this age group.

The results for middle-aged adults (30-54 years) are more concerning, as the mortality rate followed an increasing trend after 2012. For example, Figure 1(b) shows that the death rate for individuals aged 45-49 increased by around 1% per year between 2012 and 2019, making the death rate in 2019 comparable to that of 2010. See analogous results for other age strata in the 30-54 years group in Appendix A, Figures 3-7. During 2020 and 2021, the death rate for 30-54 year olds was anomalously high, reflecting the impact of the COVID-19 pandemic on this age group. Although mortality rates decreased in 2022 and 2023, they remained higher than pre-pandemic levels, consistent with previous studies^4–7^.

Previous analyses often attributed the sustained increase in mortality to the pandemic and its impact on the healthcare system and socioeconomic factors^4,6,7^. However, projecting the pre-pandemic increasing trend into the post-pandemic era suggests that the mortality rates in 2022 and 2023 might have been reached regardless of the pandemic, had the pre-pandemic increasing trend continued at the same pace (see the prediction shown by the shaded projection region in Figure 1(b)). It is likely that the high post-pandemic mortality rates have been influenced by both the factors responsible for the pre-pandemic increasing trend and the COVID-19 pandemic. The contribution of each factor cannot be disentangled with the data considered here.

The mortality rate for older adults (ages 55-89) followed a decreasing trend from 2005 to 2019, although the rate of decline slowed after 2012 (see Figure 1(c)). During the COVID-19 pandemic, the mortality rate significantly increased in 2020 and 2021. By 2022-2023, the rates relaxed to lower levels but remained higher than the levels that would have been achieved if the pre-pandemic decreasing trend had continued.

The death rate for individuals aged 90 and above shows significant variation over time (Figure 1(d)). The highest death rate was observed in 2020, likely due to COVID-19. However, this fluctuation is not a remarkable outlier compared to other years.

The observed features of mortality rates in the UK are generally reflected in individual countries (see Figures 16-30 in Appendix A for detailed results for England & Wales, Scotland, and Northern Ireland). Notably, pre-pandemic increases in mortality rates for middle-aged individuals were most pronounced in Scotland, with moderate increases observed in England & Wales. In contrast, the mortality rates for Northern Ireland exhibit significant statistical uncertainty (error bars), and an increasing mortality trend prior to the COVID-19 pandemic is not evident for middle-aged adults.

### Excess mortality

Positive excess mortality was observed in the UK before the pandemic across many age groups. However, not all these excess deaths correspond to increased mortality rates that would indicate concerning trends. Let us examine some examples from Figure 1 (with detailed plots for all age strata provided in Appendix A).

For the 1–29-year-olds, the ONS method^8^ predicts positive excess deaths between 2014 and 2016 (Figure 1(a)), likely due to an increasing population during a period when the death rate remained constant (Figure 1(e)). Conversely, for 45–49-year-olds, the positive excess deaths between 2013 and 2018 (Figure 1(b)) are likely linked to an increase in the mortality rate, reflecting a rising trend in deaths (Figure 1(f)). A pre-pandemic period with positive excess deaths occurred for 60–64-year-olds between 2013 and 2018 (Figure 1(c)). This is probably associated with an increasing population, as the mortality rate decreased during this time (Figure 1(g)).

Excess deaths were evident during the COVID-19 pandemic (2020-2021) for ages above 34 years (Figure 1(b-d) and Appendix A). Excess deaths decreased in 2022 and even became negative for some age groups over 34 in 2023 (Figure 1(b) and (c) and Appendix A). This suggests an improvement compared to the pandemic period but should not be seen as entirely positive when considering the mortality rates. On one hand, mortality rates remained significantly higher than before the pandemic for middle-aged individuals (30-54 years). On the other hand, post-pandemic death rates for older individuals (55-89 years) stayed higher than pre-pandemic levels and higher than they might have been had the pre-pandemic decreasing trend continued.

## Discussion

Our analysis reveals critical insights into age-specific mortality trends in the UK before and after the COVID-19 pandemic. We observed a concerning increase in all-cause mortality rates for middle-aged adults (30-54 years) beginning in 2012, suggesting that the current elevated rates might have occurred irrespective of the COVID-19 pandemic, due to pre-existing trends. This finding is more alarming than the British Heart Foundation’s observation of a slowdown in the decline of death rates from cardiovascular diseases since 2012 for those under 75^6^. The pooling of different age groups under 75 may have concealed potential increasing trends in mortality rates among middle-aged adults.

Our results underscore the complexity of interpreting excess mortality. While excess deaths were significant during the pandemic, reflecting its immediate impact on mortality rates, long-term trends are influenced by both changes in mortality rates and population dynamics. Consequently, excess deaths can occur during periods of constant or declining mortality rates due to population increases, as observed for some age groups in pre-pandemic periods. After the pandemic, excess mortality has relaxed and even suggests a lower-than-expected number of deaths for some age groups. This might be misleading if not properly interpreted since mortality rates are not lower than before the pandemic. In general, long-term trends in excess mortality may be of limited value on their own and should be interpreted in conjunction with population trends and mortality rates.

Another complication for excess mortality estimates is that they depend on the specific choice of the baseline used to describe “normal” conditions (i.e., conditions without extraordinary events such as COVID-19). There is also a risk for sophisticated models to be used as a “black box” by practitioners who may lack a thorough understanding of the underlying assumptions and limitations. Moreover, their complexity may undermine public trust^10^.

Given the complexity of interpreting excess deaths, we suggest that simpler measures like mortality rates remain valuable for public health practitioners due to their ease of estimation and interpretation. In general, the combined use of multiple measures provides a more comprehensive understanding of mortality patterns, enabling better-informed public health responses and policy decisions.

In summary, our study emphasises the importance of recognising pre-pandemic mortality trends and their interplay with the COVID-19 pandemic, as well as their influence on current health outcomes. This highlights the need for targeted interventions and enhanced healthcare planning to effectively manage current and future public health challenges. These insights are not only relevant to the UK but also hold significance for other countries experiencing similar mortality trends.

## Data Availability

All data produced in the present work are contained in the manuscript

## Data availability

Population and mortality data used in our analyses were collated in Ref. ^12^.

## Appendix A. Supplementary figures

Supplementary figures related to this article are provided in Appendix A.

## Acknowledgements

Funding support is acknowledged from the UKRI COVID-19 Longitudinal Health and Wellbeing National Core Study, a Medical Research Council Fellowship (MR/W021455/1) and a Research Leave Award.

