## Appendix A for "Age-Specific All-Cause Mortality Trends in the UK: Pre-Pandemic Increases and the Complex Impact of COVID-19"

### Appendix A. Supplementary figures

This document provides figures for all the age strata considered in the work. The contents are structured in two sections: one presenting yearly statistics for the whole UK, and the other providing plots of yearly mortality rates by country.

##### Contents


1. Yearly mortality rates, excess mortality, population and deaths in the UK
    - Less than 1 year
    - 1 to 29 years
    - 30 to 34 years
    - 35 to 39 years
    - 40 to 44 years
    - 45 to 49 years
    - 50 to 54 years
    - 55 to 59 years
    - 60 to 64 years
    - 65 to 69 years
    - 70 to 74 years
    - 75 to 79 years
    - 80 to 84 years
    - 85 to 89 years
    - 90 years and above
2. Mortality rates by country
    - Less than 1 year
    - 1 to 29 years
    - 30 to 34 years
    - 35 to 39 years
    - 40 to 44 years
    - 45 to 49 years
    - 50 to 54 years
    - 55 to 59 years
    - 60 to 64 years
    - 65 to 69 years
    - 70 to 74 years
    - 75 to 79 years
    - 80 to 84 years
    - 85 to 89 years
    - 90 years and above

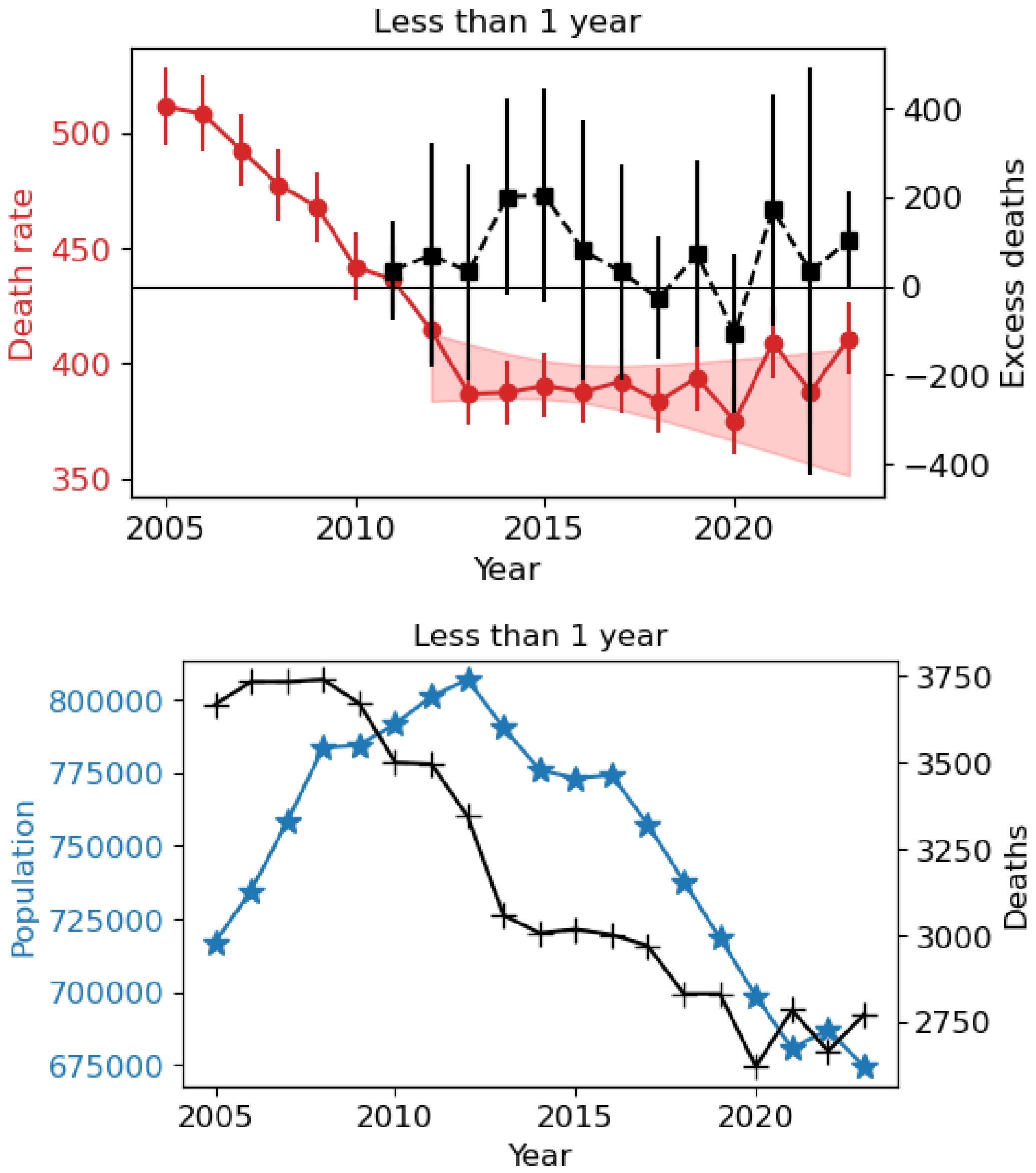


**Figure 1:** Yearly mortality statistics for **less than 1-year-olds** in the UK. Upper panel: Mortality rates (red circles) and excess mortality (black squares). The shaded red region represents the 95% confidence interval for a linear fit to the mortality rate from 2012 to 2019. The extended region for 2020-2023 predicts the death rate assuming pre-pandemic trends had continued. Lower panel: Population (blue stars) and number of deaths (black crosses).

Back to List of Figures

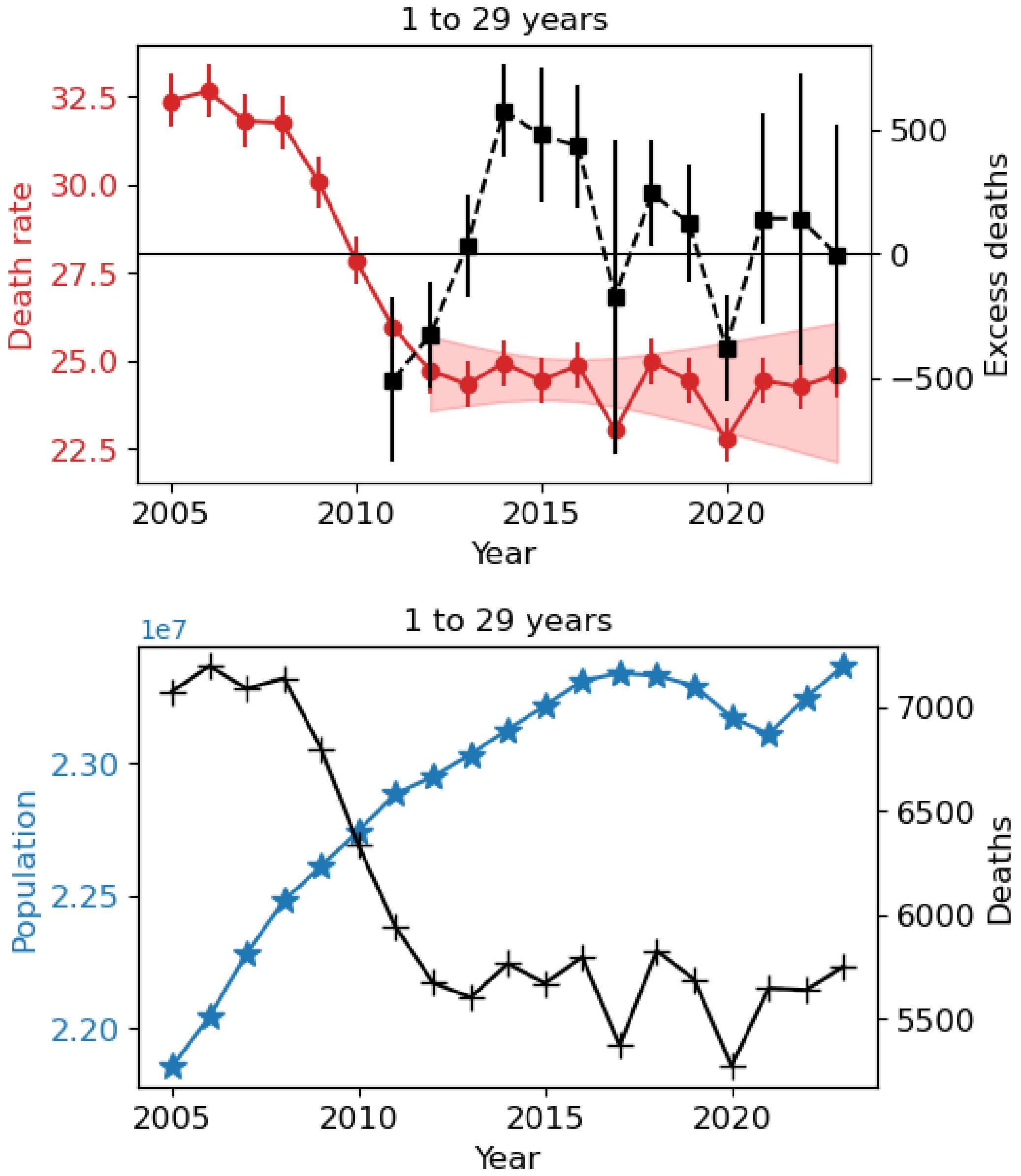


**Figure 2:** Yearly mortality statistics for **1 to 29-year-olds** in the UK. Upper panel: Mortality rates (red circles) and excess mortality (black squares). The shaded red region represents the 95% confidence interval for a linear fit to the mortality rate from 2012 to 2019. The extended region for 2020-2023 predicts the death rate assuming pre-pandemic trends had continued. Lower panel: Population (blue stars) and number of deaths (black crosses).

Back to List of Figures

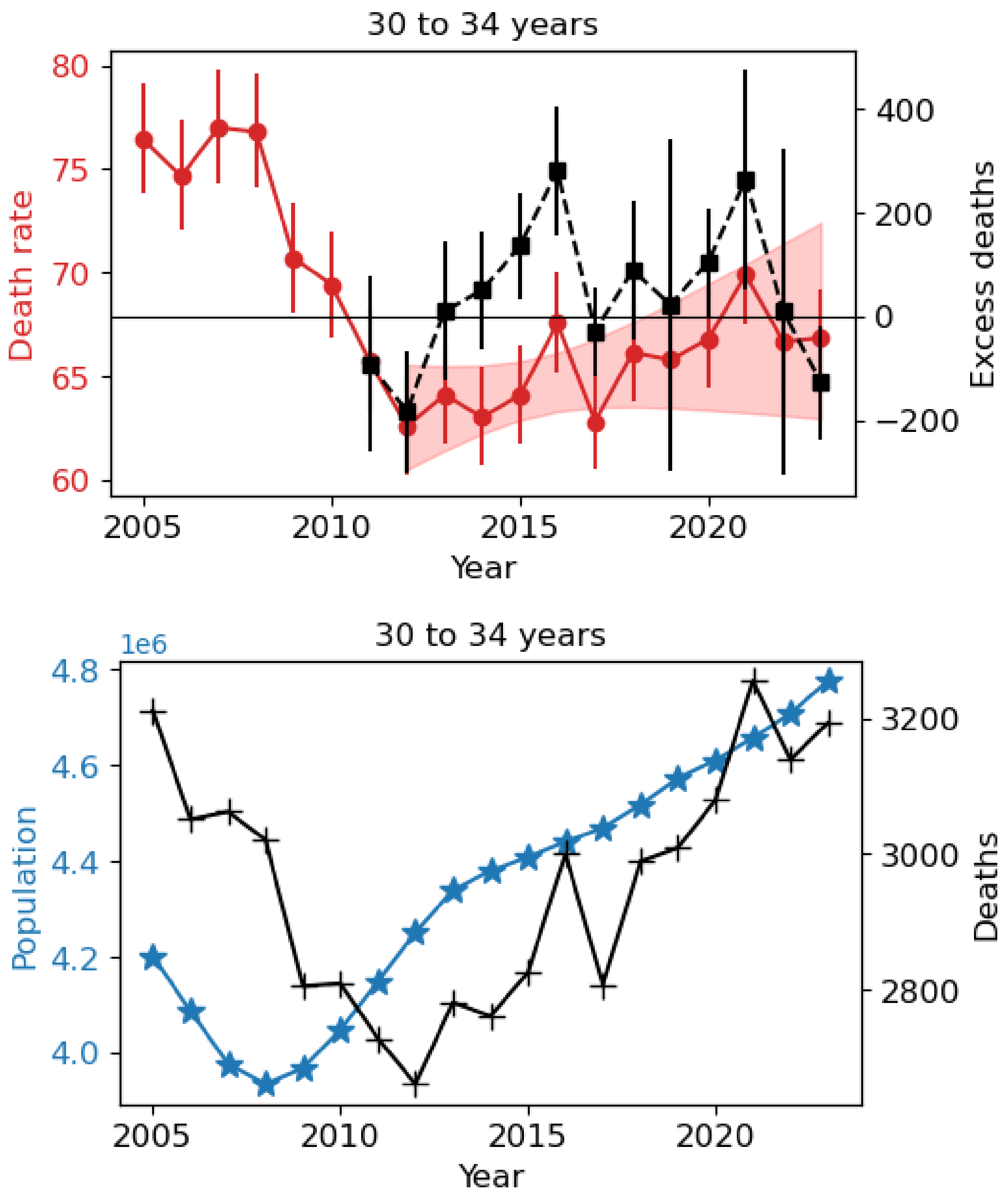


**Figure 3:** Yearly mortality statistics for **30 to 34-year-olds** in the UK. Upper panel: Mortality rates (red circles) and excess mortality (black squares). The shaded red region represents the 95% confidence interval for a linear fit to the mortality rate from 2012 to 2019. The extended region for 2020-2023 predicts the death rate assuming pre-pandemic trends had continued. Lower panel: Population (blue stars) and number of deaths (black crosses).

Back to List of Figures

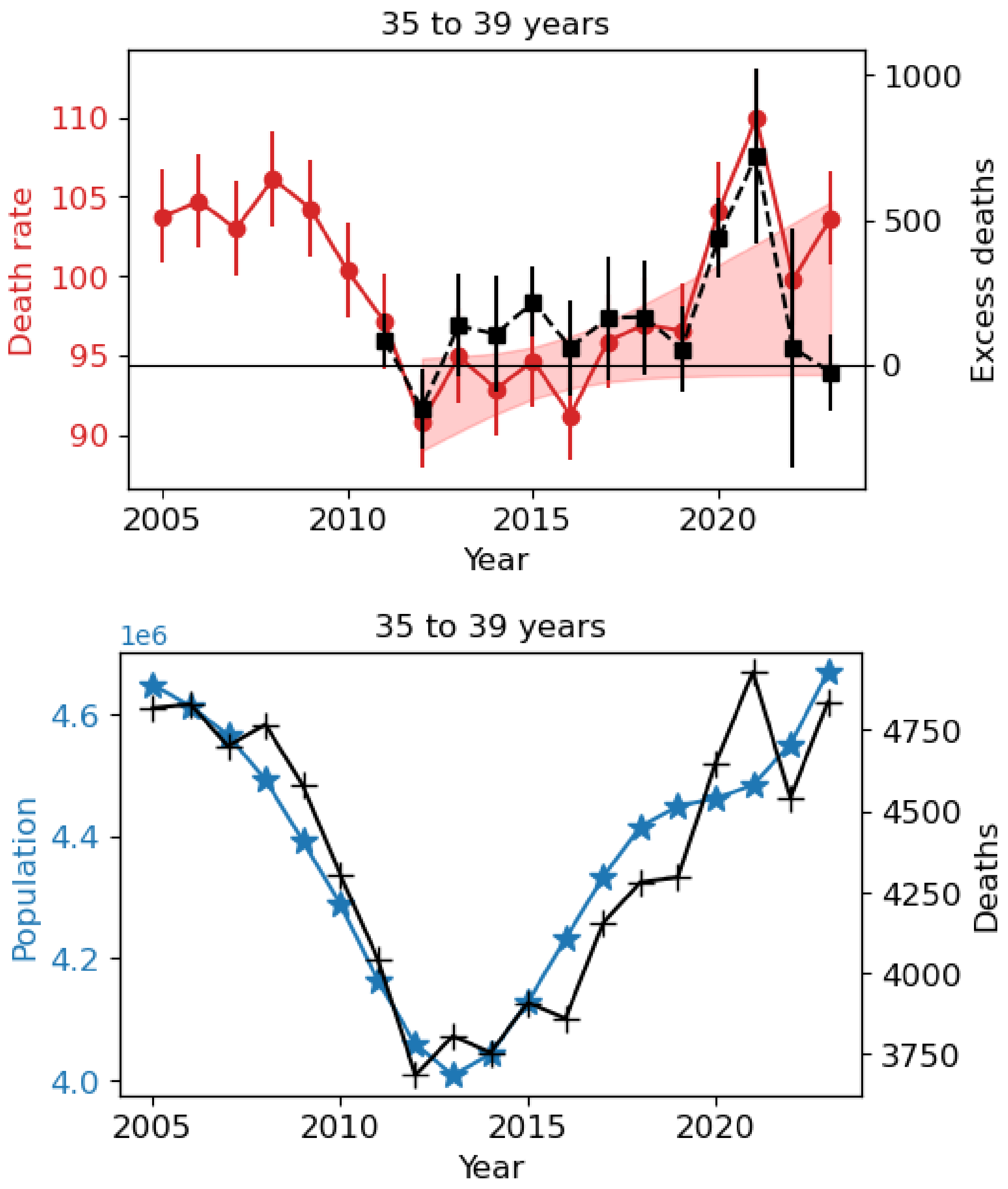


**Figure 4:** Yearly mortality statistics for **35 to 39-year-olds** in the UK. Upper panel: Mortality rates (red circles) and excess mortality (black squares). The shaded red region represents the 95% confidence interval for a linear fit to the mortality rate from 2012 to 2019. The extended region for 2020-2023 predicts the death rate assuming pre-pandemic trends had continued. Lower panel: Population (blue stars) and number of deaths (black crosses).

Back to List of Figures

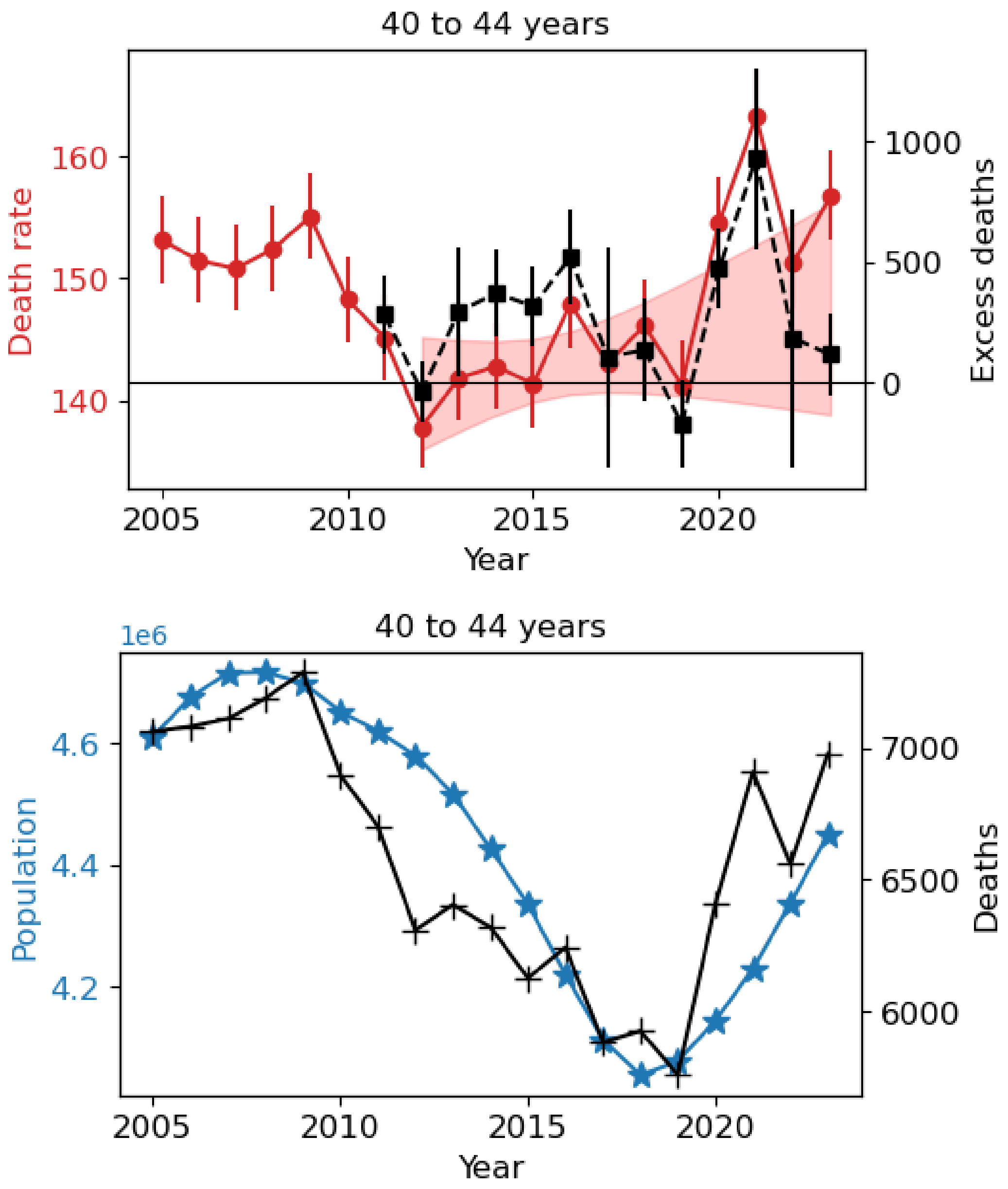


**Figure 5:** Yearly mortality statistics for **40 to 45-year-olds** in the UK. Upper panel: Mortality rates (red circles) and excess mortality (black squares). The shaded red region represents the 95% confidence interval for a linear fit to the mortality rate from 2012 to 2019. The extended region for 2020-2023 predicts the death rate assuming pre-pandemic trends had continued. Lower panel: Population (blue stars) and number of deaths (black crosses).

Back to List of Figures

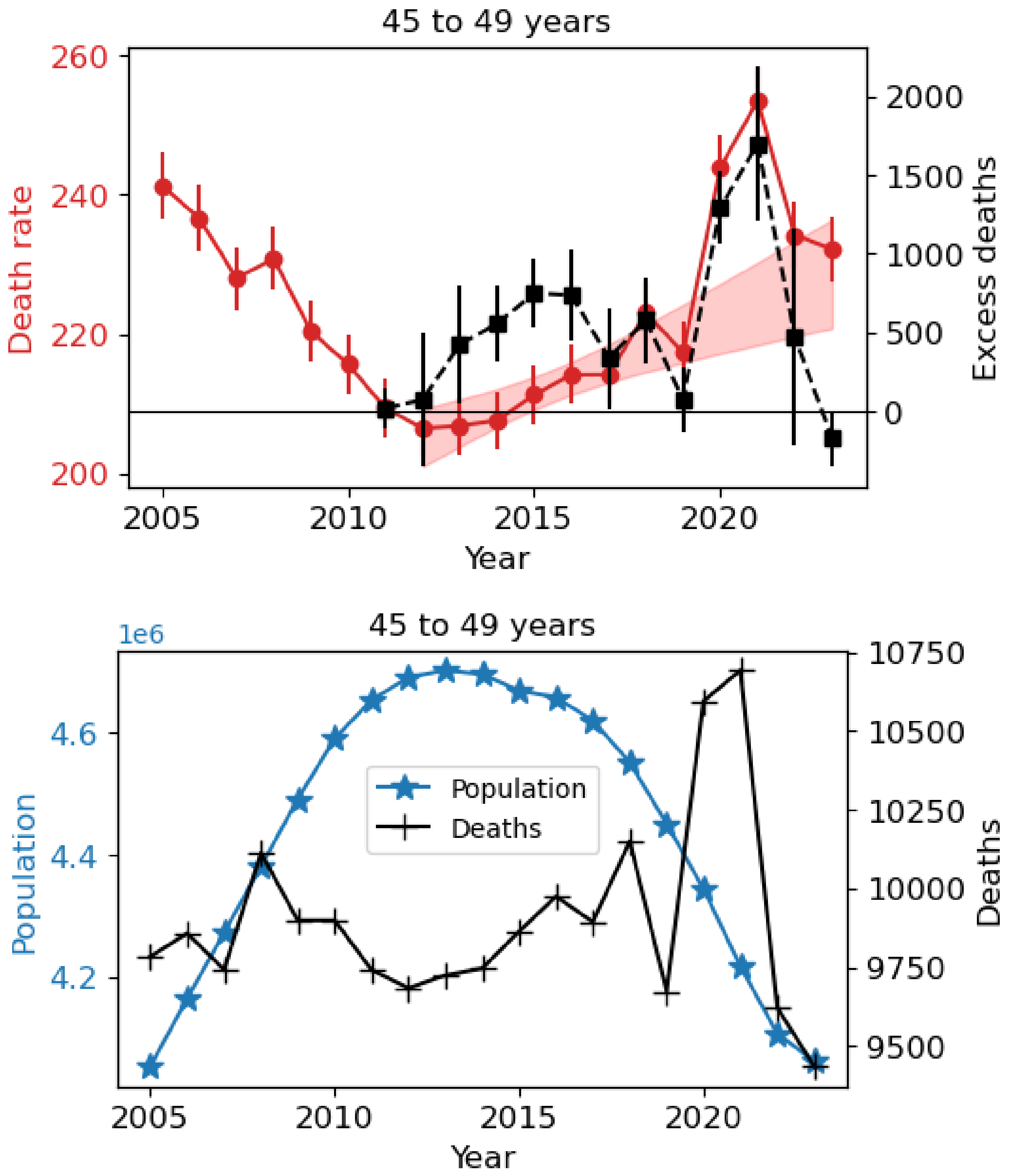


**Figure 6:** Yearly mortality statistics for **45 to 49-year-olds** in the UK. Upper panel: Mortality rates (red circles) and excess mortality (black squares). The shaded red region represents the 95% confidence interval for a linear fit to the mortality rate from 2012 to 2019. The extended region for 2020-2023 predicts the death rate assuming pre-pandemic trends had continued. Lower panel: Population (blue stars) and number of deaths (black crosses).

Back to List of Figures

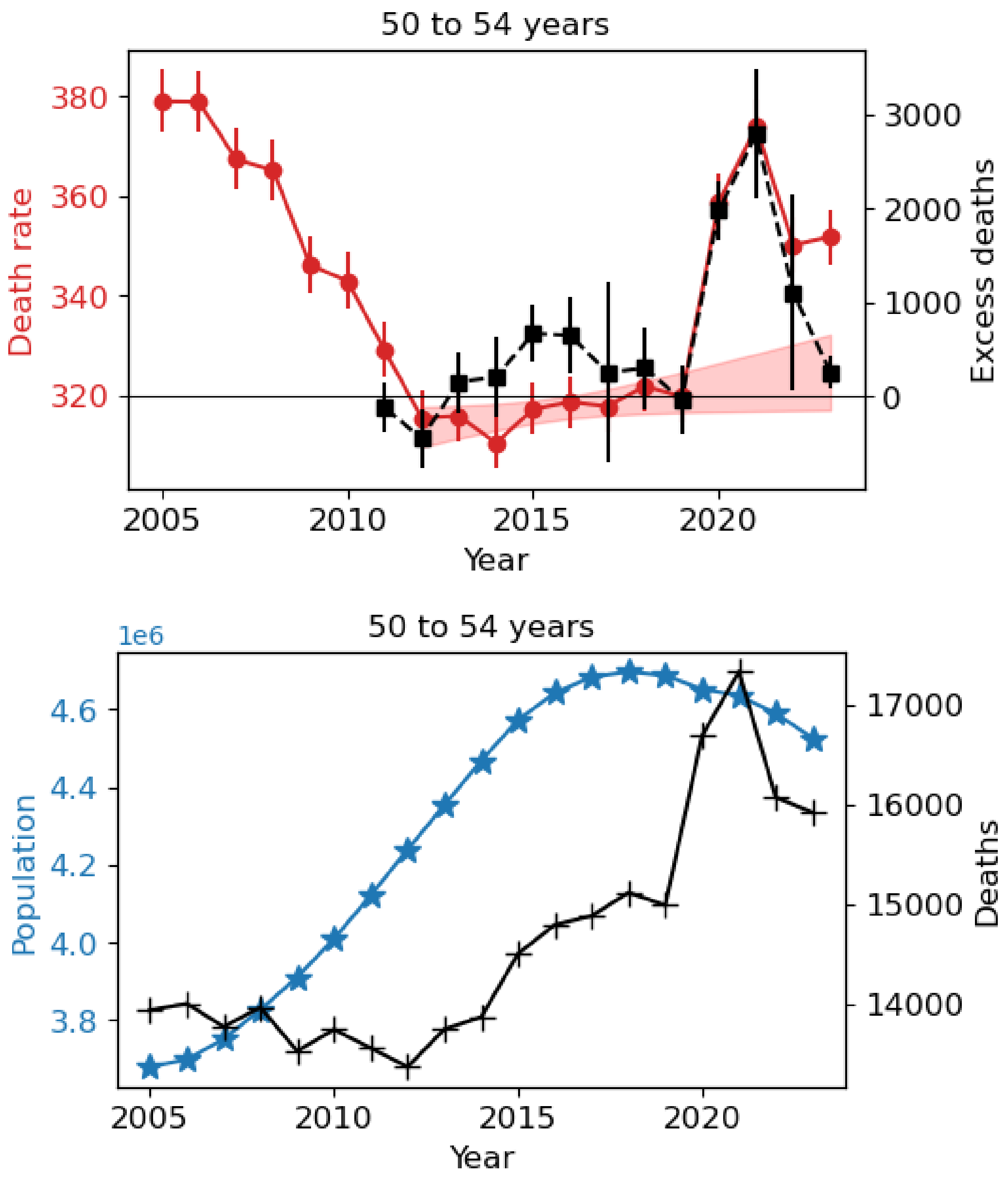


**Figure 7:** Yearly mortality statistics for **50 to 54-year-olds** in the UK. Upper panel: Mortality rates (red circles) and excess mortality (black squares). The shaded red region represents the 95% confidence interval for a linear fit to the mortality rate from 2012 to 2019. The extended region for 2020-2023 predicts the death rate assuming pre-pandemic trends had continued. Lower panel: Population (blue stars) and number of deaths (black crosses).

Back to List of Figures

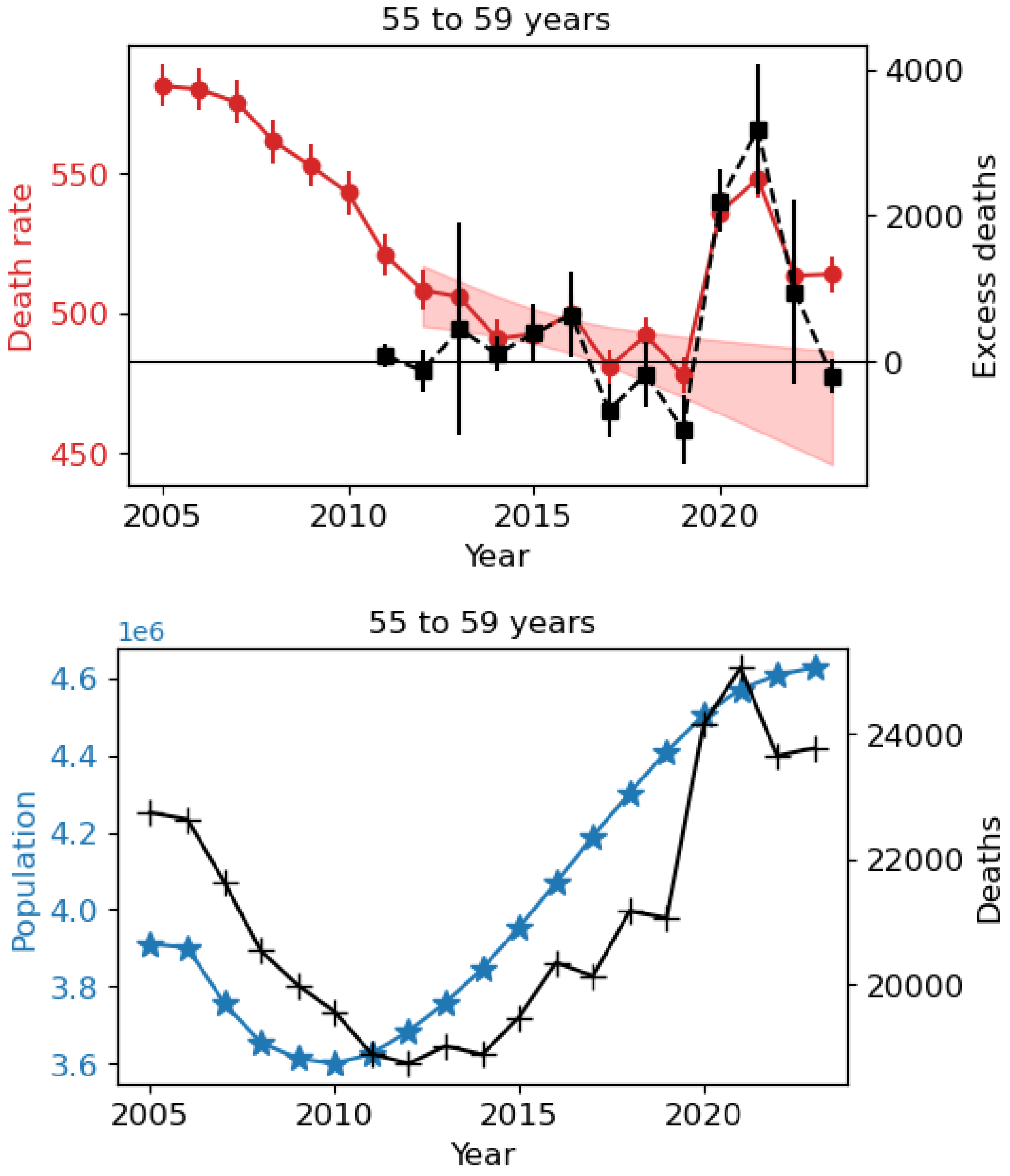


**Figure 8:** Yearly mortality statistics for **55 to 59-year-olds** in the UK. Upper panel: Mortality rates (red circles) and excess mortality (black squares). The shaded red region represents the 95% confidence interval for a linear fit to the mortality rate from 2012 to 2019. The extended region for 2020-2023 predicts the death rate assuming pre-pandemic trends had continued. Lower panel: Population (blue stars) and number of deaths (black crosses).

Back to List of Figures

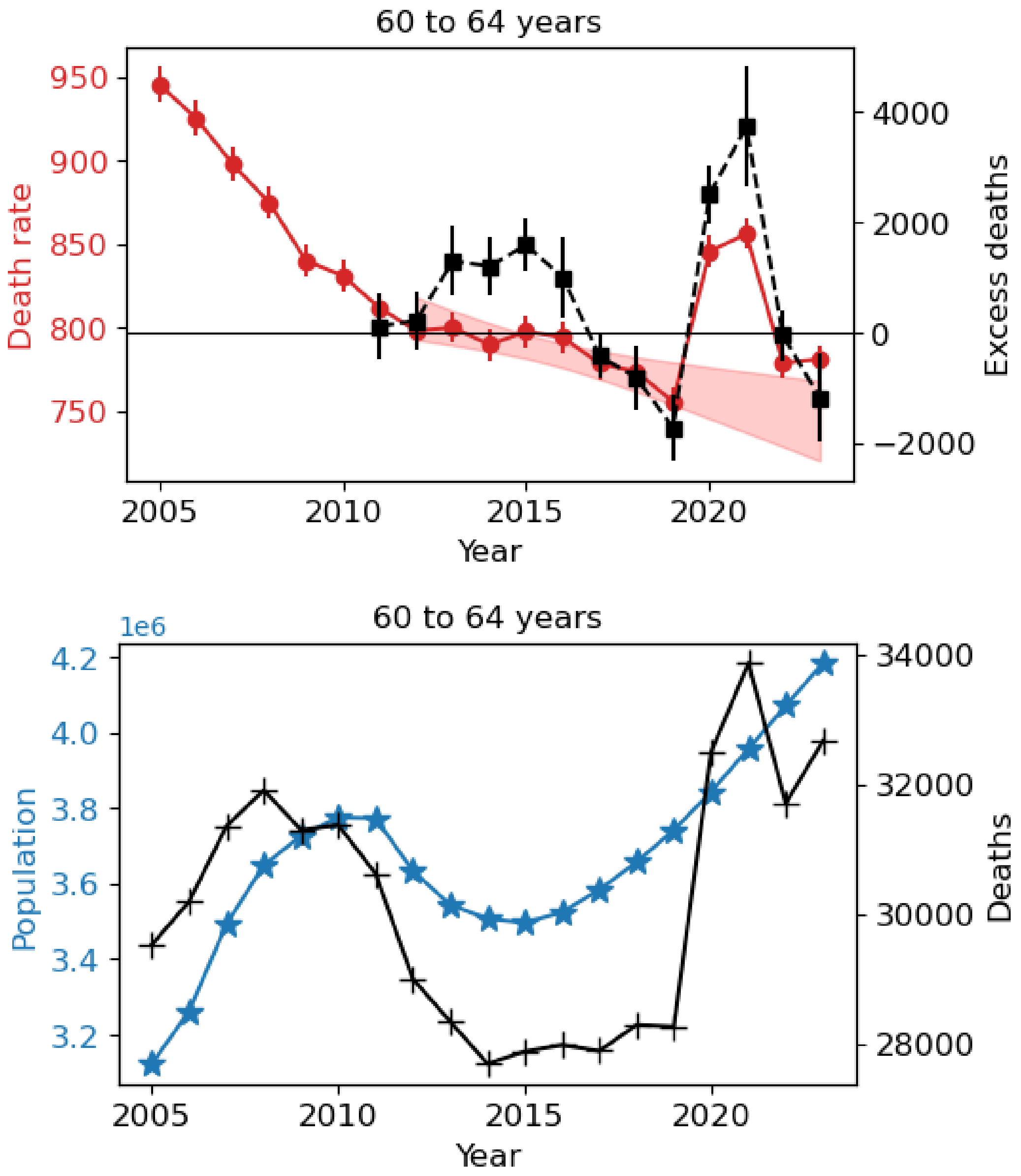


**Figure 9:** Yearly mortality statistics for **60 to 64-year-olds** in the UK. Upper panel: Mortality rates (red circles) and excess mortality (black squares). The shaded red region represents the 95% confidence interval for a linear fit to the mortality rate from 2012 to 2019. The extended region for 2020-2023 predicts the death rate assuming pre-pandemic trends had continued. Lower panel: Population (blue stars) and number of deaths (black crosses).

Back to List of Figures

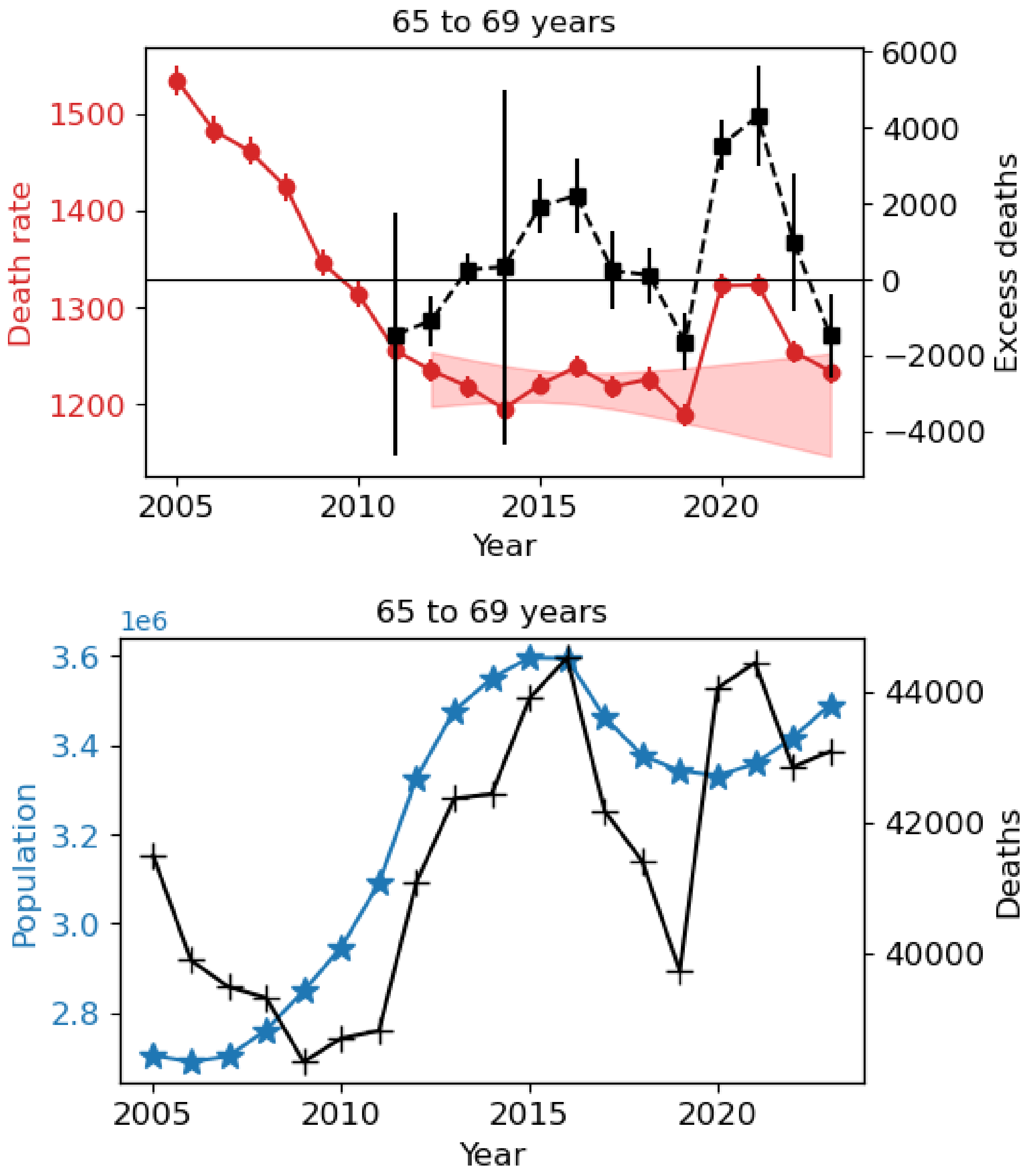


**Figure 10:** Yearly mortality statistics for **65 to 69-year-olds** in the UK. Upper panel: Mortality rates (red circles) and excess mortality (black squares). The shaded red region represents the 95% confidence interval for a linear fit to the mortality rate from 2012 to 2019. The extended region for 2020-2023 predicts the death rate assuming pre-pandemic trends had continued. Lower panel: Population (blue stars) and number of deaths (black crosses).

Back to List of Figures

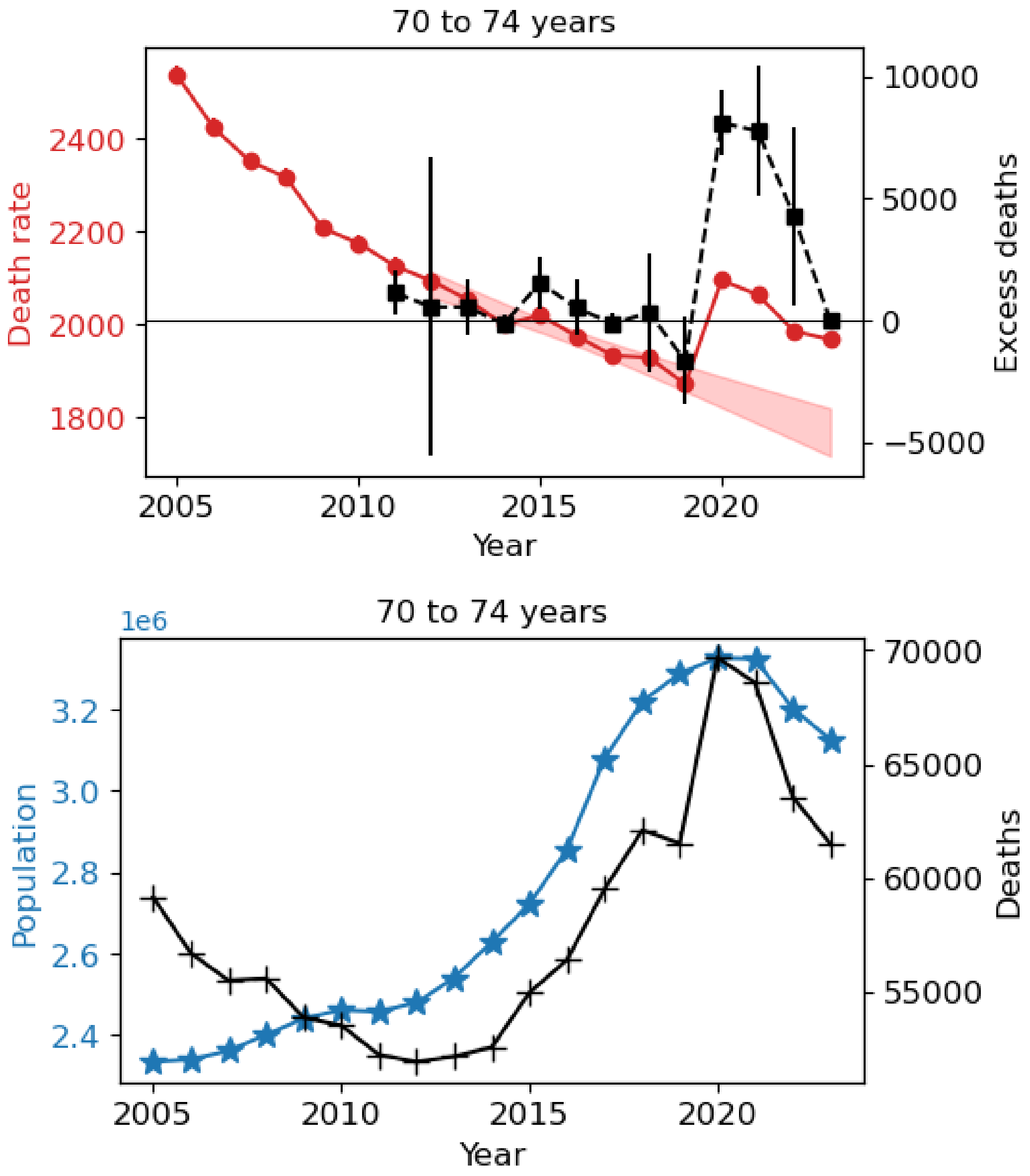


**Figure 11:** Yearly mortality statistics for **70 to 74-year-olds** in the UK. Upper panel: Mortality rates (red circles) and excess mortality (black squares). The shaded red region represents the 95% confidence interval for a linear fit to the mortality rate from 2012 to 2019. The extended region for 2020-2023 predicts the death rate assuming pre-pandemic trends had continued. Lower panel: Population (blue stars) and number of deaths (black crosses).

Back to List of Figures

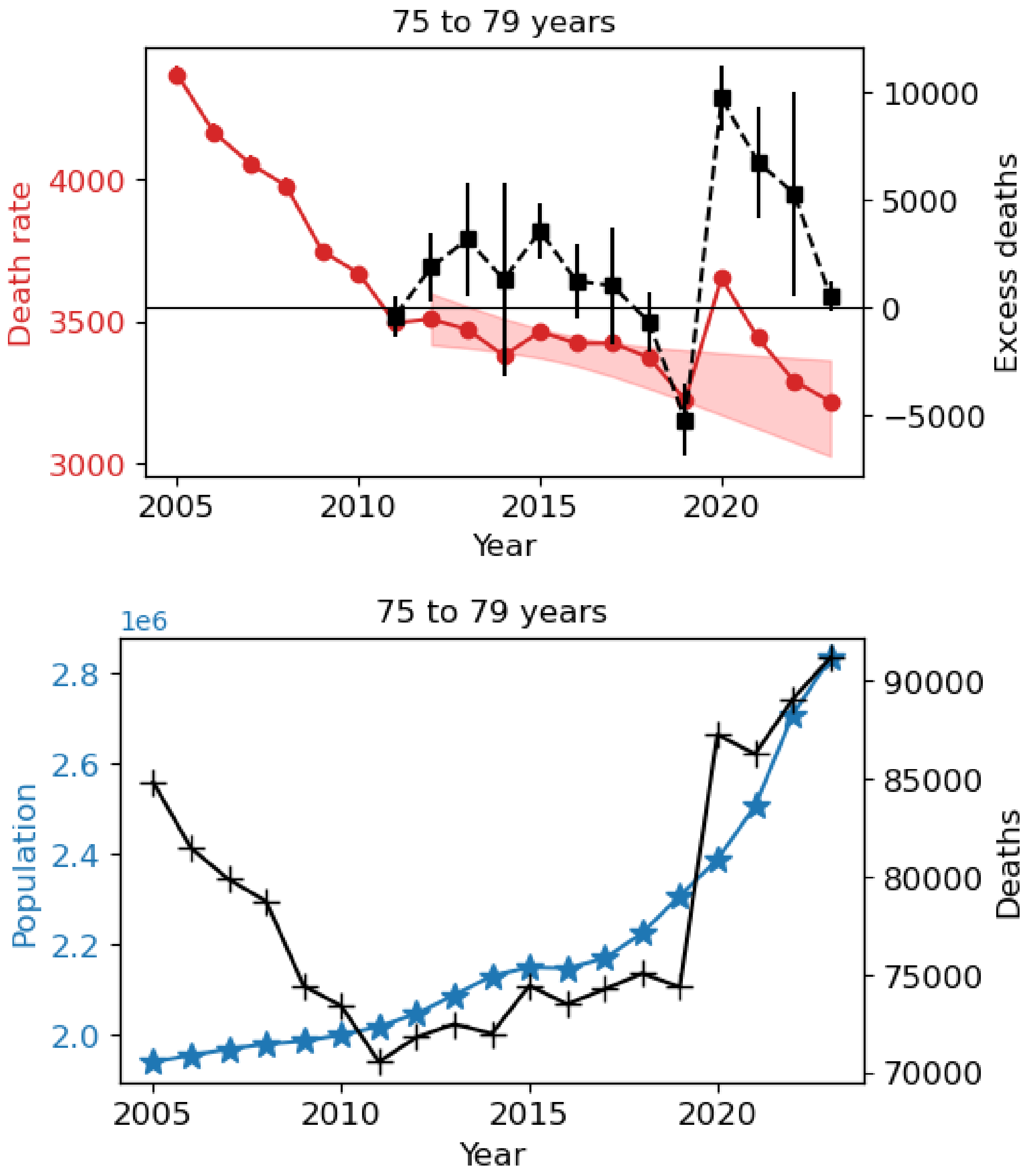


**Figure 12:** Yearly mortality statistics for **75 to 79-year-olds** in the UK. Upper panel: Mortality rates (red circles) and excess mortality (black squares). The shaded red region represents the 95% confidence interval for a linear fit to the mortality rate from 2012 to 2019. The extended region for 2020-2023 predicts the death rate assuming pre-pandemic trends had continued. Lower panel: Population (blue stars) and number of deaths (black crosses).

Back to List of Figures

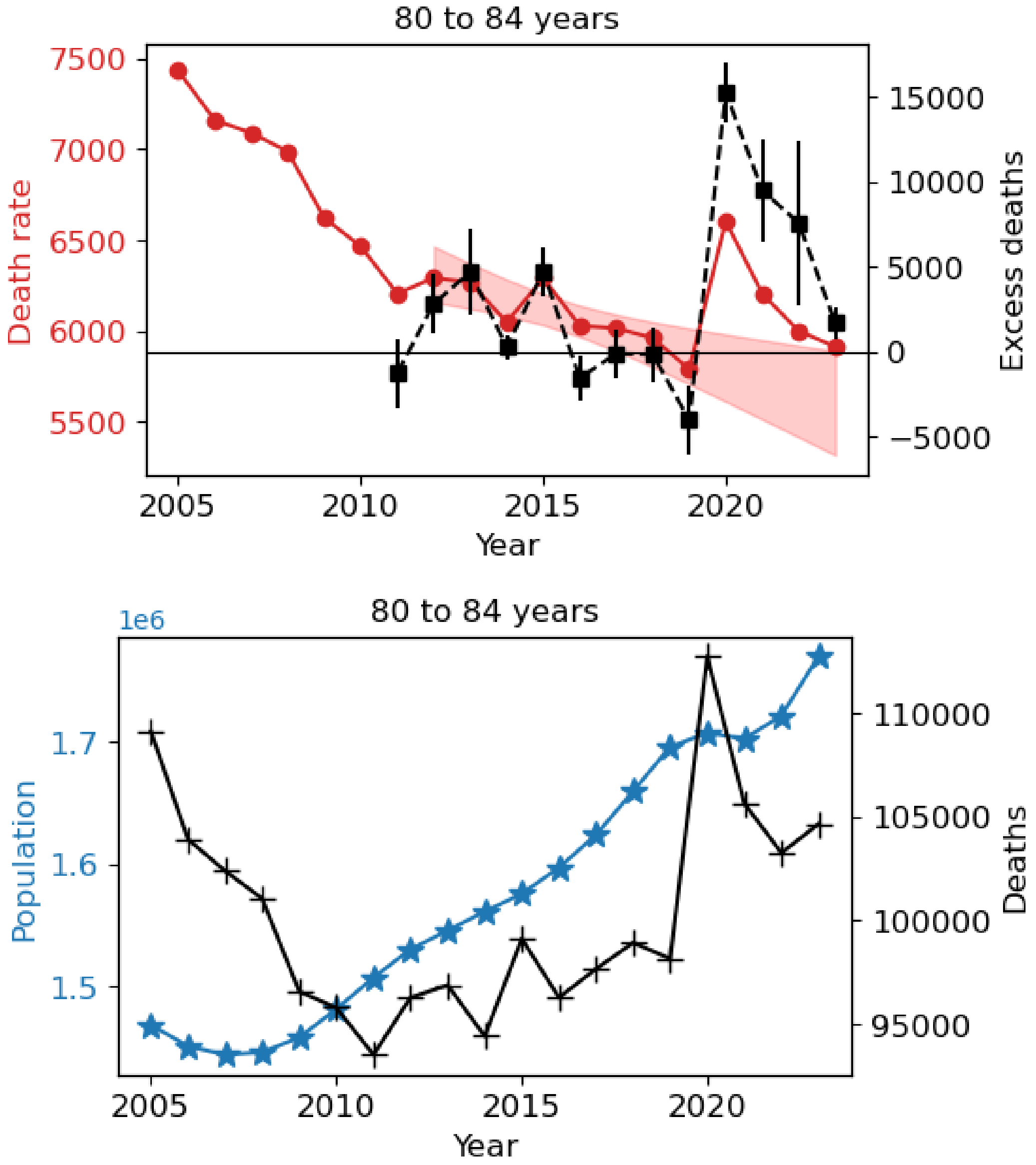


**Figure 13:** Yearly mortality statistics for **80 to 84-year-olds** in the UK. Upper panel: Mortality rates (red circles) and excess mortality (black squares). The shaded red region represents the 95% confidence interval for a linear fit to the mortality rate from 2012 to 2019. The extended region for 2020-2023 predicts the death rate assuming pre-pandemic trends had continued. Lower panel: Population (blue stars) and number of deaths (black crosses).

Back to List of Figures

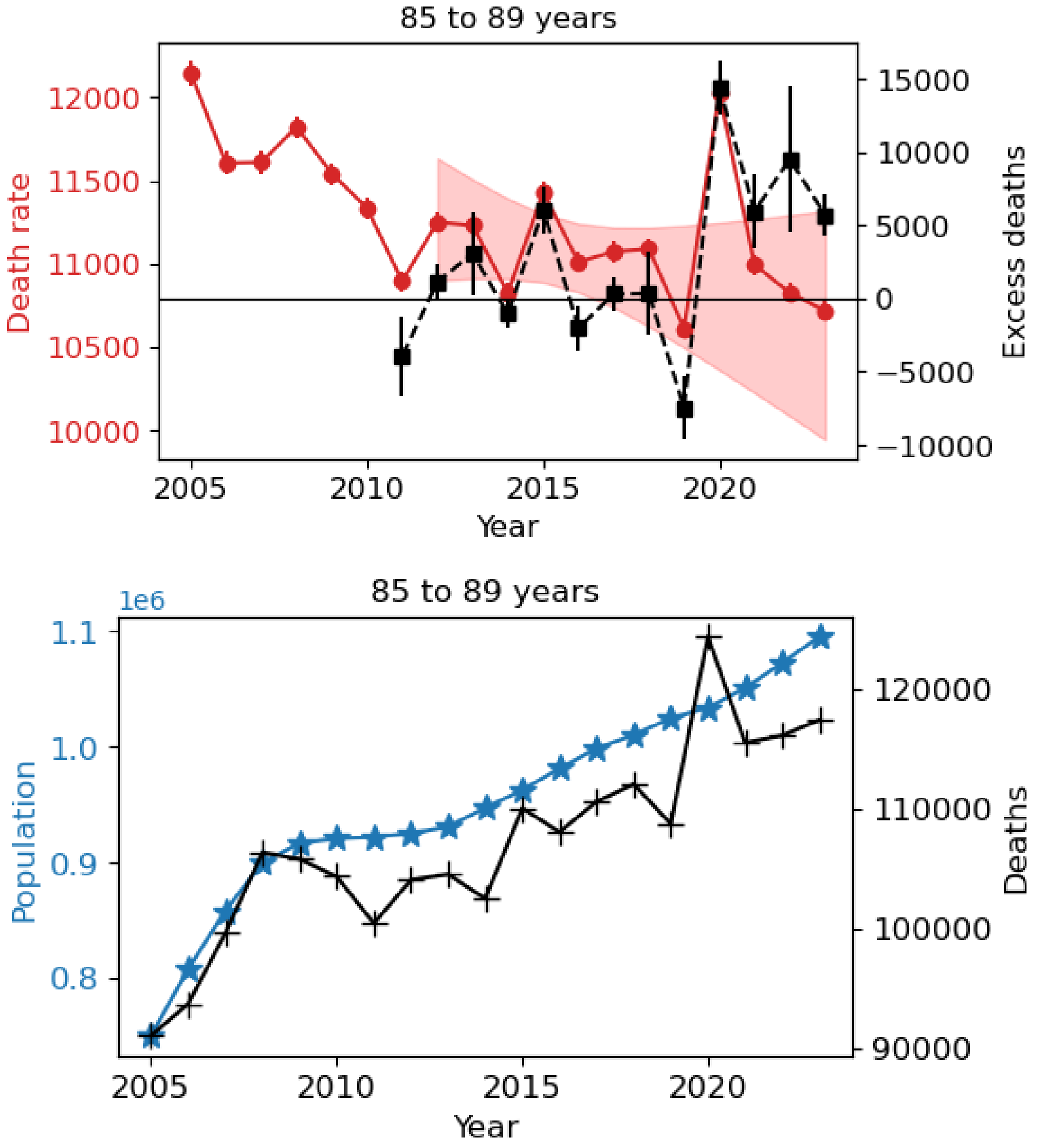


**Figure 14:** Yearly mortality statistics for **85 to 89-year-olds** in the UK. Upper panel: Mortality rates (red circles) and excess mortality (black squares). The shaded red region represents the 95% confidence interval for a linear fit to the mortality rate from 2012 to 2019. The extended region for 2020-2023 predicts the death rate assuming pre-pandemic trends had continued. Lower panel: Population (blue stars) and number of deaths (black crosses).

Back to List of Figures

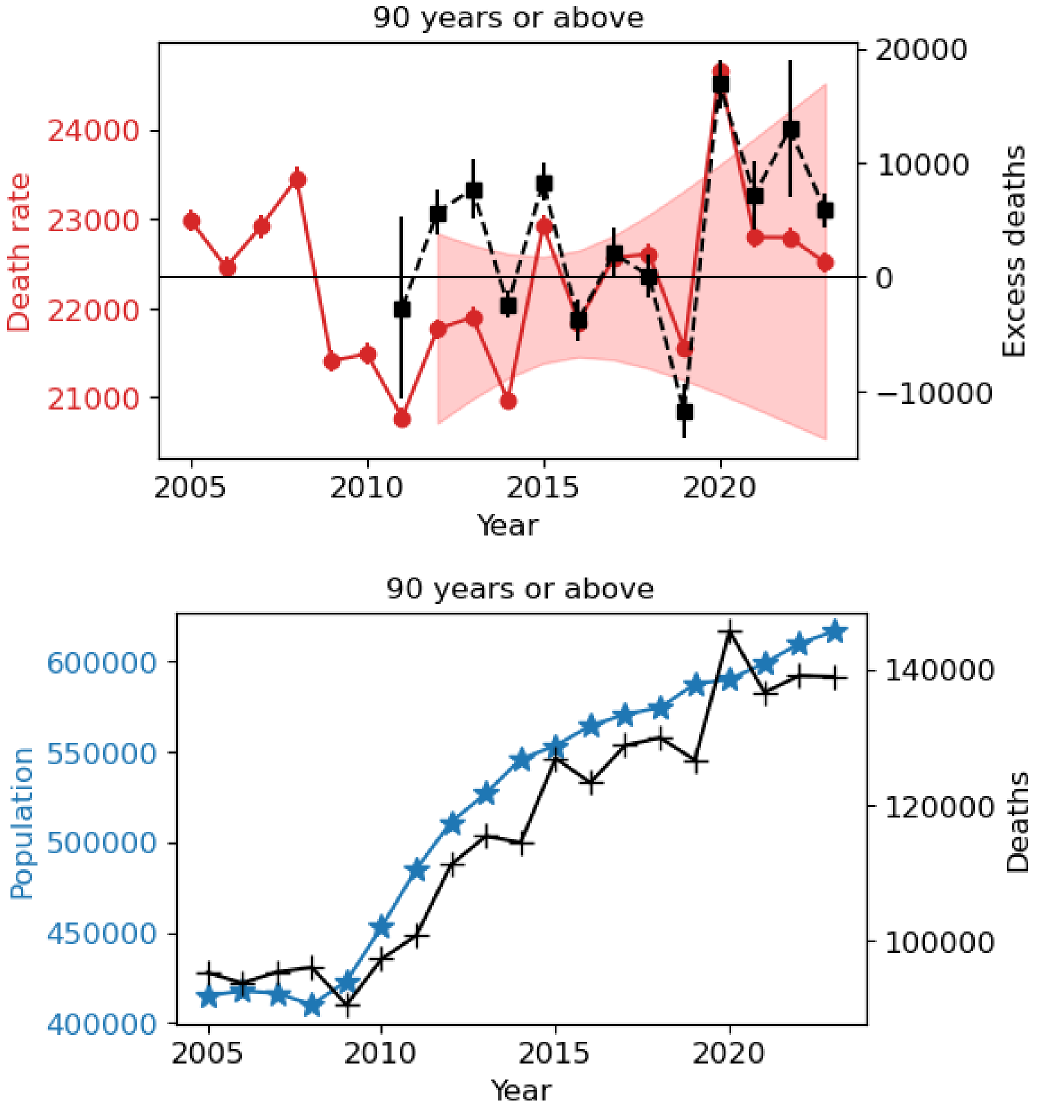


**Figure 15:** Yearly mortality statistics for **90 year-olds and above** in the UK. Upper panel: Mortality rates (red circles) and excess mortality (black squares). The shaded red region represents the 95% confidence interval for a linear fit to the mortality rate from 2012 to 2019. The extended region for 2020-2023 predicts the death rate assuming pre-pandemic trends had continued. Lower panel: Population (blue stars) and number of deaths (black crosses).

Back to List of Figures

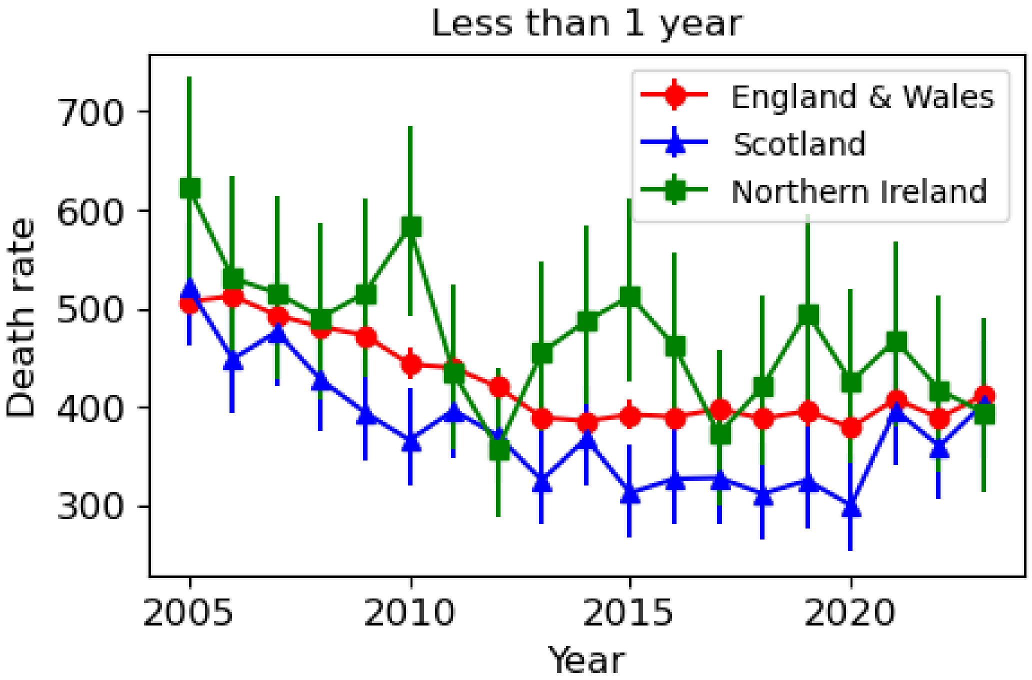


**Figure 16:** Yearly mortality rate for **less than 1-year-olds** in England & Wales (red circles), Scotland (blue triangles) and Northern Ireland (green squares).

Back to List of Figures

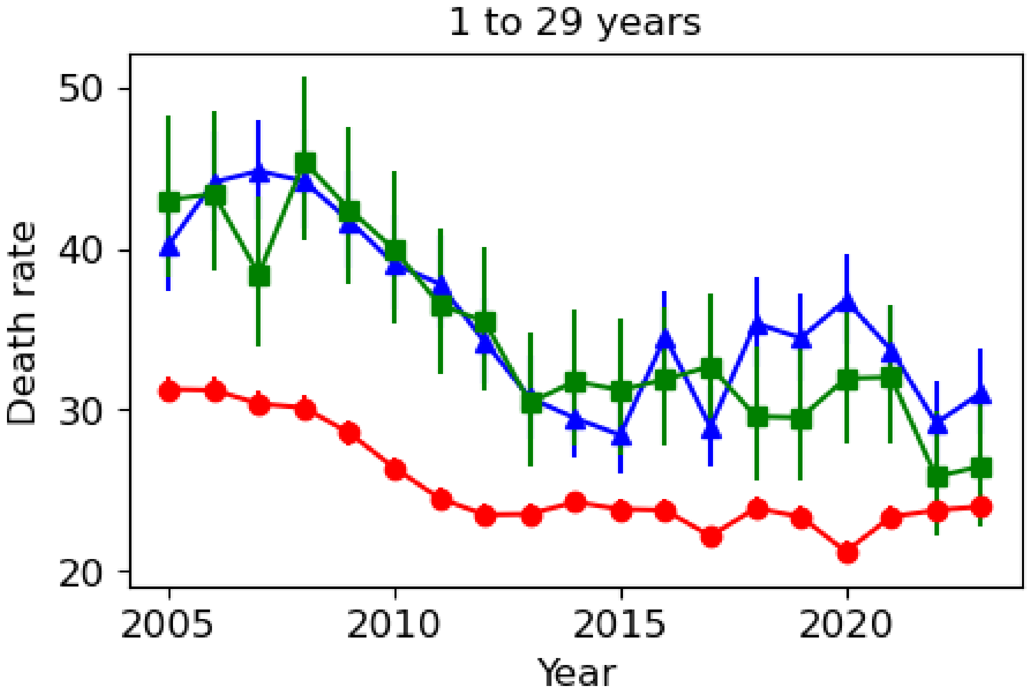


**Figure 17:** Yearly mortality rate for **1 to 29-year-olds** in England & Wales (red circles), Scotland (blue triangles) and Northern Ireland (green squares).

Back to List of Figures

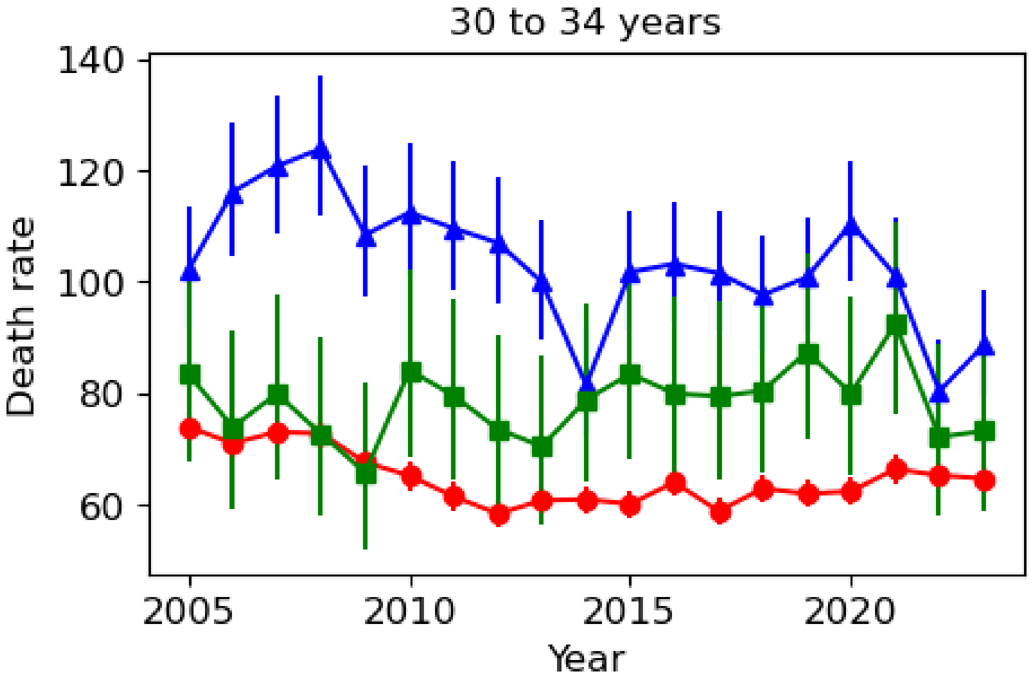


**Figure 18:** Yearly mortality rate for **30 to 34-year-olds** in England & Wales (red circles), Scotland (blue triangles) and Northern Ireland (green squares).

Back to List of Figures

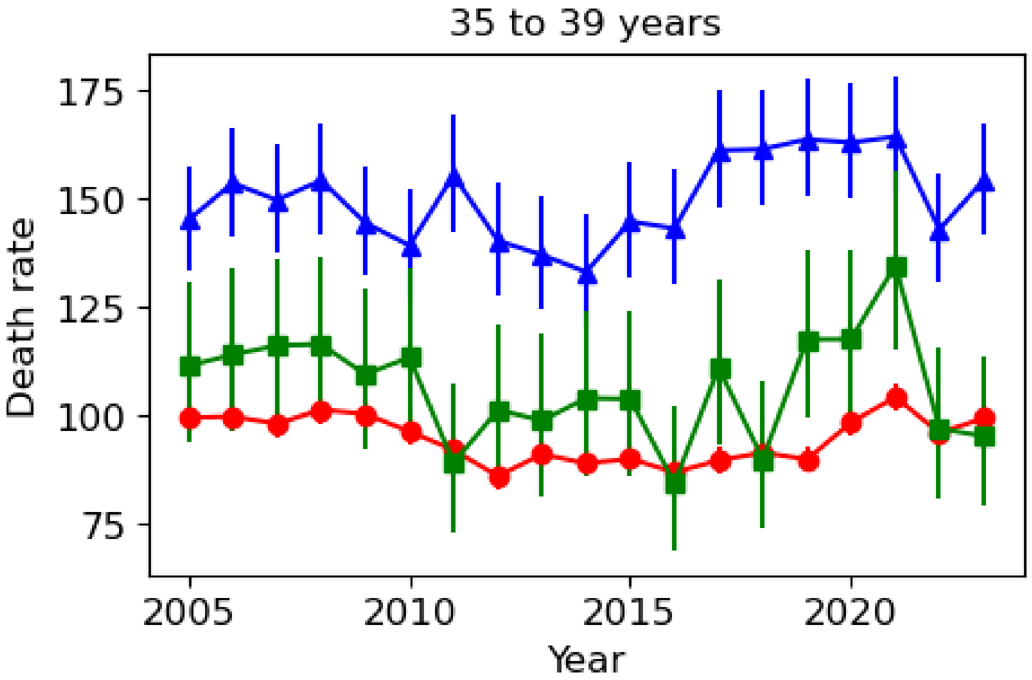


**Figure 19:** Yearly mortality rate for **35 to 39-year-olds** in England & Wales (red circles), Scotland (blue triangles) and Northern Ireland (green squares).

Back to List of Figures

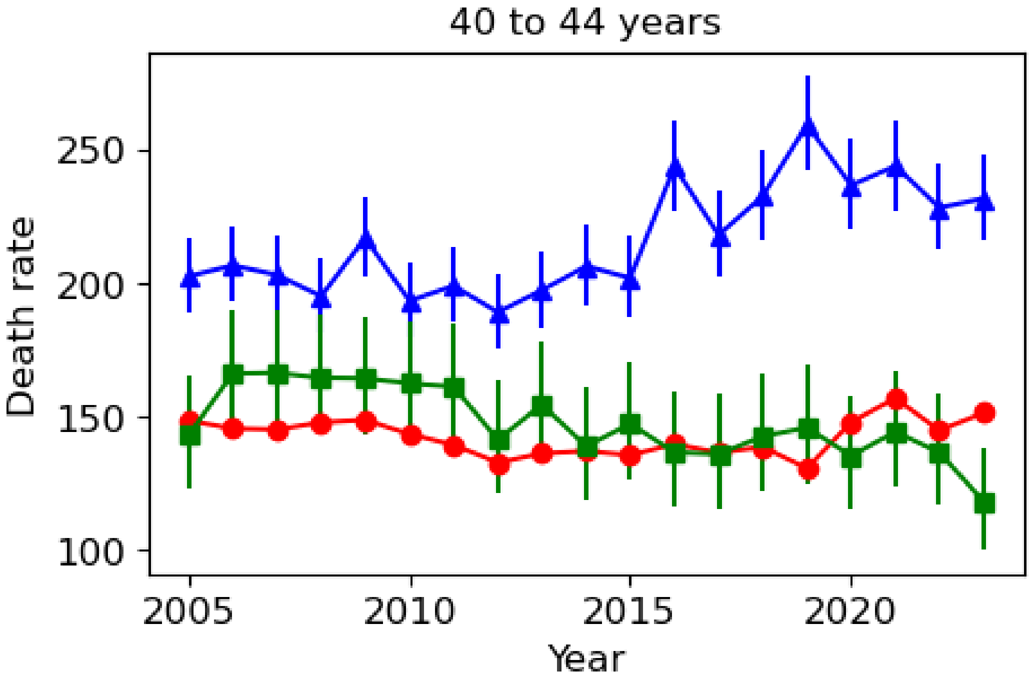


**Figure 20:** Yearly mortality rate for **40 to 44-year-olds** in England & Wales (red circles), Scotland (blue triangles) and Northern Ireland (green squares).

Back to List of Figures

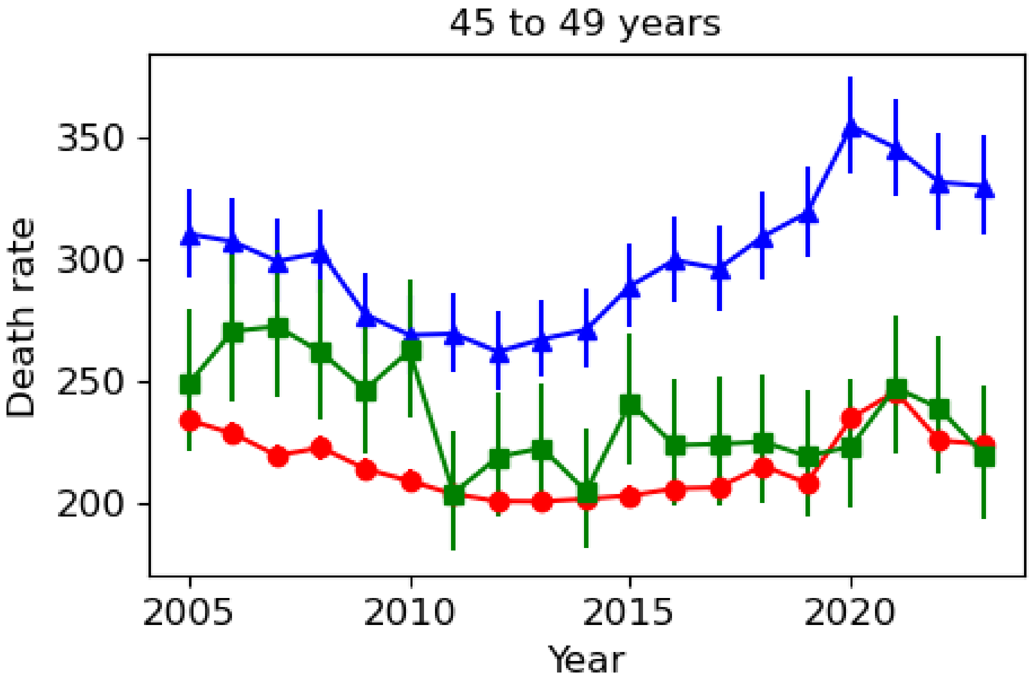


**Figure 21:** Yearly mortality rate for **35 to 39-year-olds** in England & Wales (red circles), Scotland (blue triangles) and Northern Ireland (green squares).

Back to List of Figures

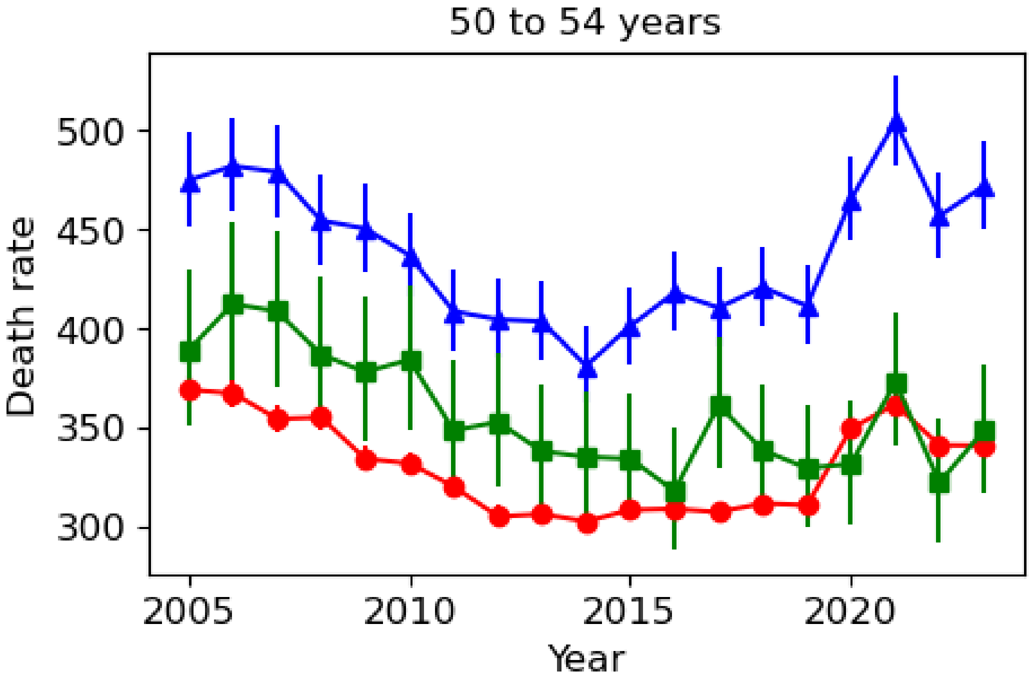


**Figure 22:** Yearly mortality rate for **50 to 54-year-olds** in England & Wales (red circles), Scotland (blue triangles) and Northern Ireland (green squares).

Back to List of Figures

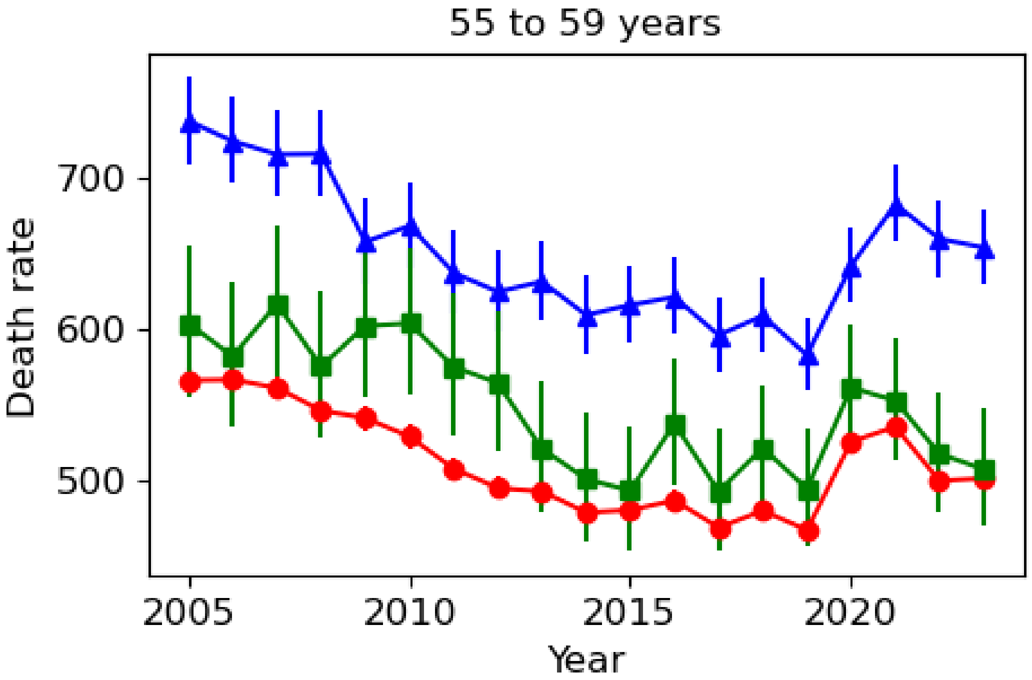


**Figure 23:** Yearly mortality rate for **55 to 59-year-olds** in England & Wales (red circles), Scotland (blue triangles) and Northern Ireland (green squares).

Back to List of Figures

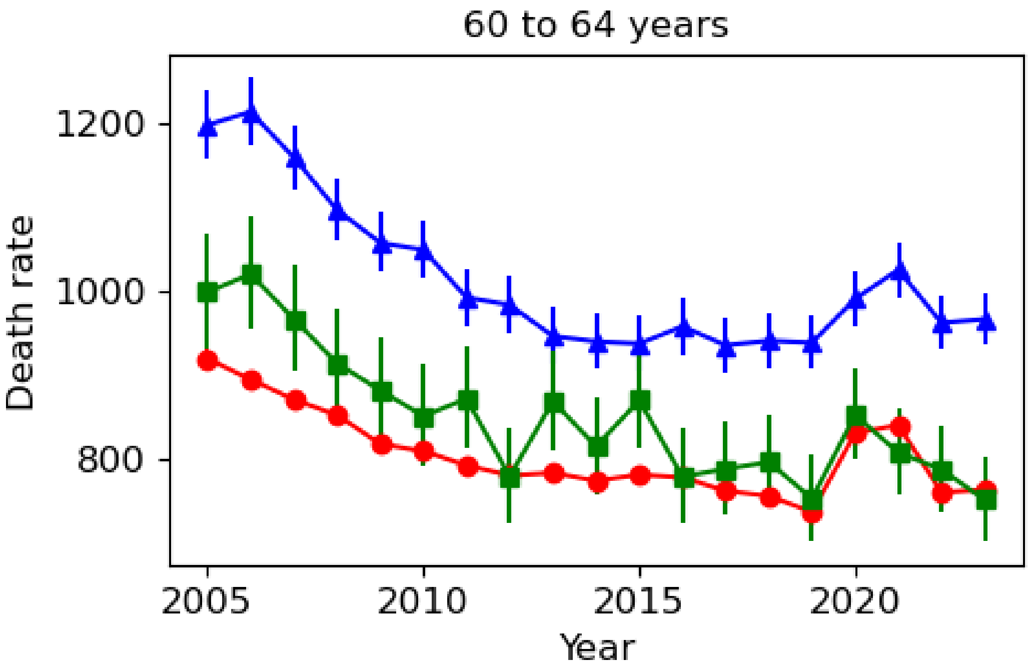


**Figure 24:** Yearly mortality rate for **60 to 64-year-olds** in England & Wales (red circles), Scotland (blue triangles) and Northern Ireland (green squares).

Back to List of Figures

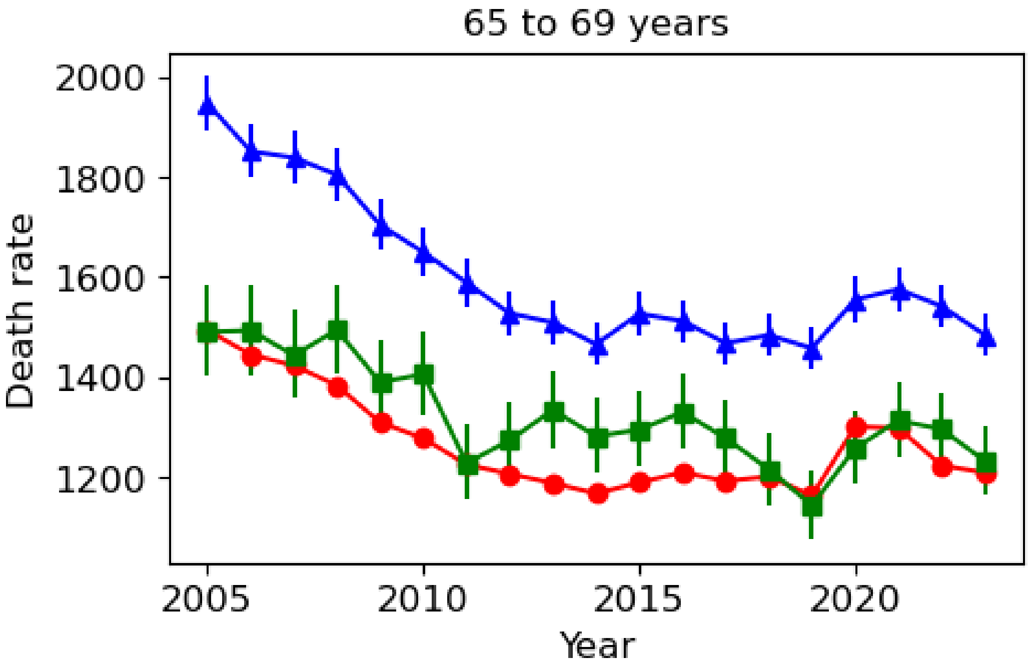


**Figure 25:** Yearly mortality rate for **65 to 69-year-olds** in England & Wales (red circles), Scotland (blue triangles) and Northern Ireland (green squares).

Back to List of Figures

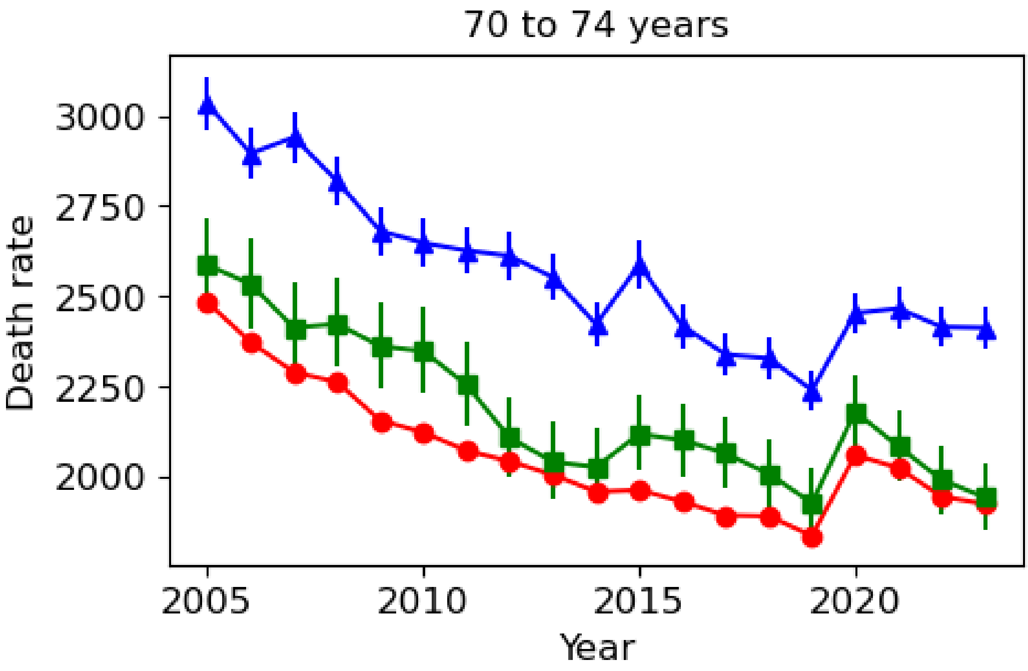


**Figure 26:** Yearly mortality rate for **70 to 74-year-olds** in England & Wales (red circles), Scotland (blue triangles) and Northern Ireland (green squares).

Back to List of Figures

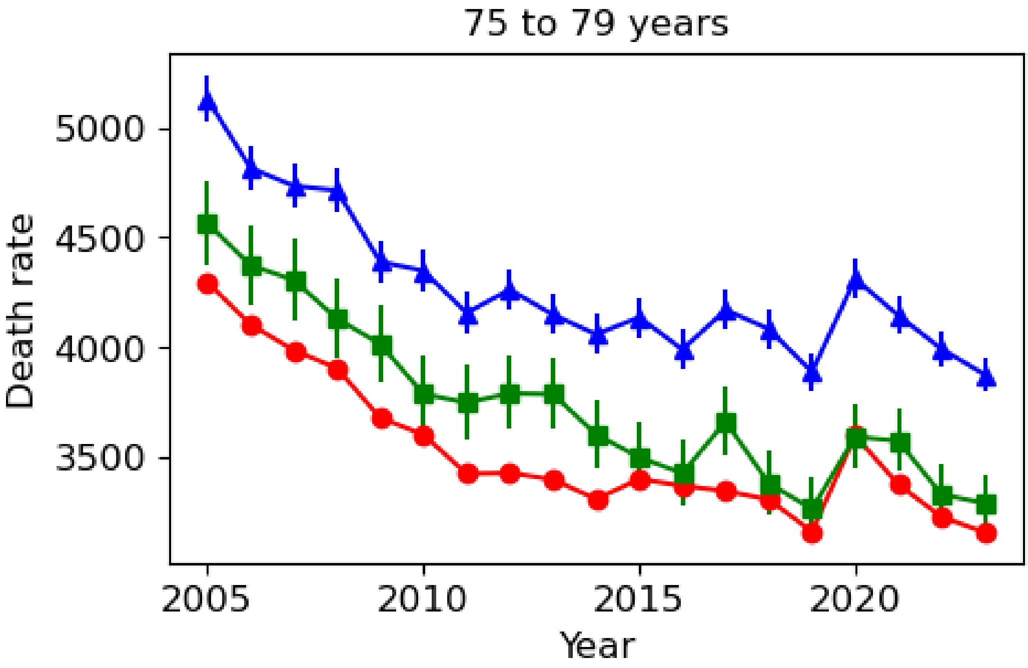


**Figure 27:** Yearly mortality rate for **75 to 79-year-olds** in England & Wales (red circles), Scotland (blue triangles) and Northern Ireland (green squares).

Back to List of Figures

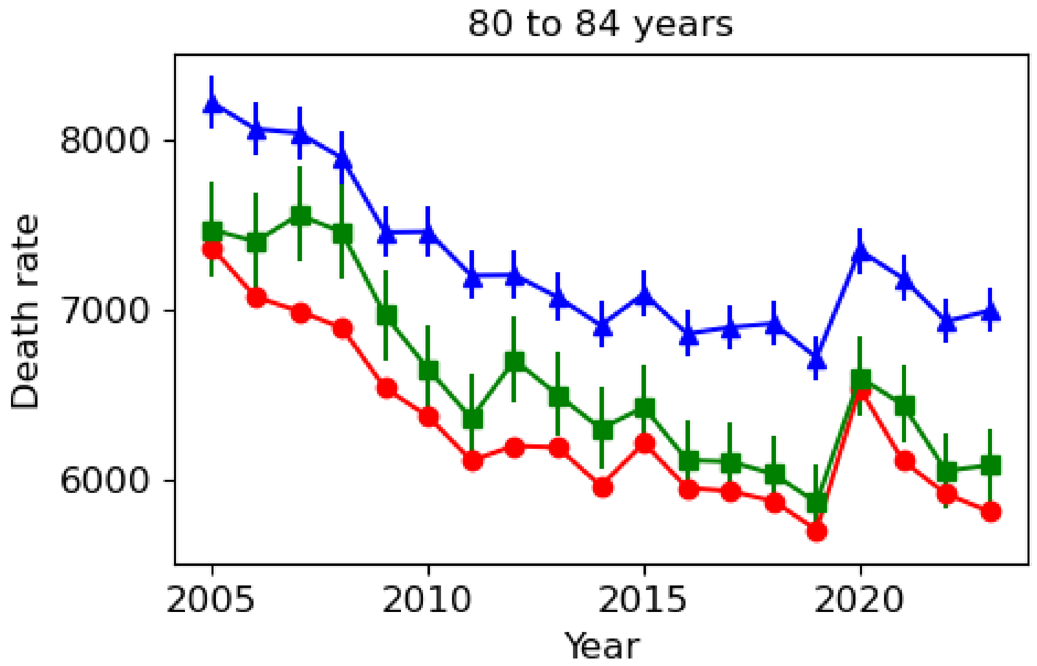


**Figure 28:** Yearly mortality rate for **80 to 84-year-olds** in England & Wales (red circles), Scotland (blue triangles) and Northern Ireland (green squares).

Back to List of Figures

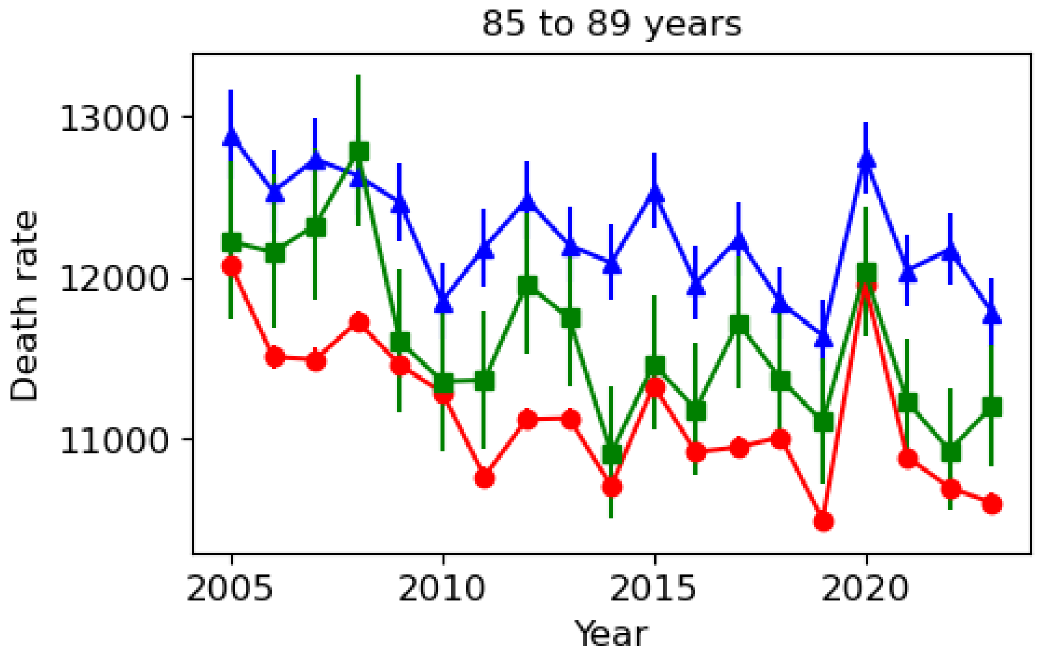


**Figure 29:** Yearly mortality rate for **85 to 89-year-olds** in England & Wales (red circles), Scotland (blue triangles) and Northern Ireland (green squares).

Back to List of Figures

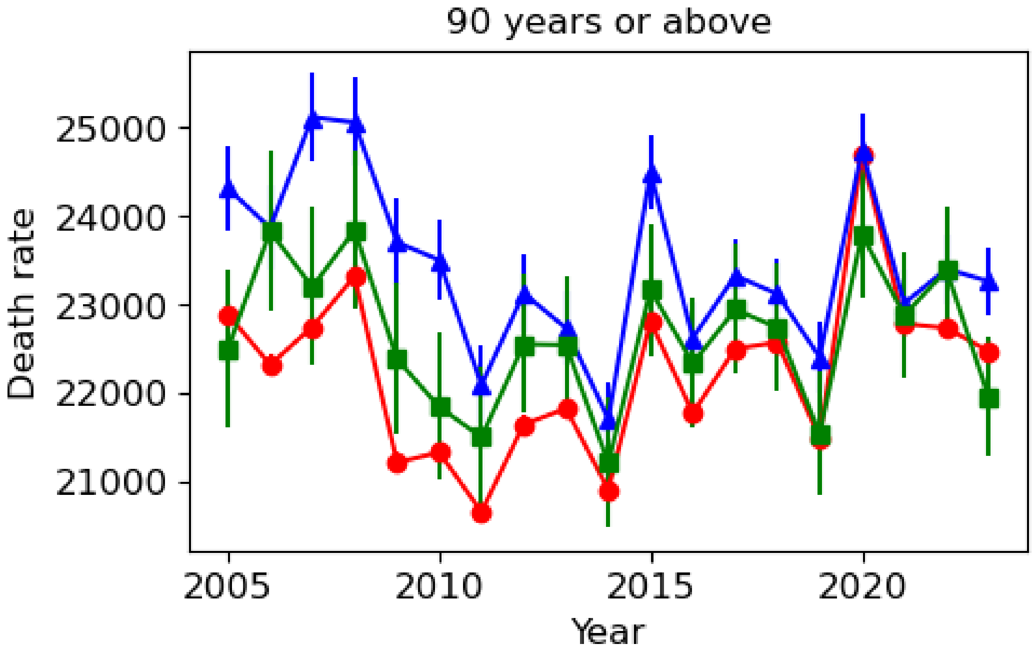


**Figure 30:** Yearly mortality rate for **90 year-olds and above** in England & Wales (red circles), Scotland (blue triangles) and Northern Ireland (green squares).

Back to List of Figures
